# Post-Initiation Care Patterns and Weight Loss With Semaglutide and Tirzepatide for Obesity in Clinical Practice

**DOI:** 10.64898/2026.10.04.26364715

**Authors:** Huanhuan Yang, Chungsoo Kim, Joseph S. Ross, Chenxi Huang, Kyungseon Choi, Bo Kang, Adith S. Arun, Huilin Tang, Yong Chen, Hua Xu, Mona Sharifi, Zhihui Li, Harlan M. Krumholz, Yuan Lu

## Abstract

**Background:** Randomized trials show substantial weight loss with semaglutide and tirzepatide under structured care, but little is known about whether patients in routine practice continue therapy, reach a maintenance dose, and have their weight measured.

**Objectives:** To describe the first year after initiation of semaglutide or tirzepatide for obesity, including care patterns, maintenance-dose attainment, weight documentation, and total body weight loss (TBWL).

**Methods:** This retrospective cohort study used Epic Cosmos, an electronic health record network of >350 US health systems, to identify adults with obesity and without diabetes who initiated injectable semaglutide or tirzepatide in 2024. Outcomes over 12 months included prescribing persistence, discontinuation, switching, add-on therapy, and reinitiation; attainment of Food and Drug Administration (FDA)-labeled maintenance doses; availability of follow-up weight; and TBWL across progressively restricted populations.

**Results:** Among 743,153 adults (60.1% semaglutide; mean age, 48.5 years; 72.5% female), 12-month prescribing persistence was 29.9% for semaglutide and 35.9% for tirzepatide. FDA-labeled maintenance-dose attainment was 28.3% and 55.0%, respectively, and only 8.8% and 16.4% remained prescribing persistent at a maintenance dose with a recorded weight at 12 months. About half of initiators lacked a 12-month weight. Among all initiators, ≥5% TBWL was documented in 25.6% (semaglutide) and 30.4% (tirzepatide), rising to 51.9% and 61.9% among those with recorded weight, 66.1% and 79.5% among those also prescribing persistent, and 82.9% and 87.3% among those further at a maintenance dose.

**Conclusions:** In routine practice, most patients initiating semaglutide or tirzepatide did not remain on therapy, reach a maintenance dose, and have weight documented through the first year. Documented weight loss was greatest among those completing each step of care, suggesting continuation, dose escalation, and monitoring as targets for improvement.

**CENTRAL ILLUSTRATION:** Care Cascade and Weight-Loss Outcomes Among Semaglutide and Tirzepatide Initiators, Standardized to 100 Initiators.

Numbers are per 100 initiators and were rounded so that categories at each step sum to the preceding step. At the 12-month dose step, prescribed dose was missing for 1 per 100 semaglutide initiators and <1 per 100 tirzepatide initiators.

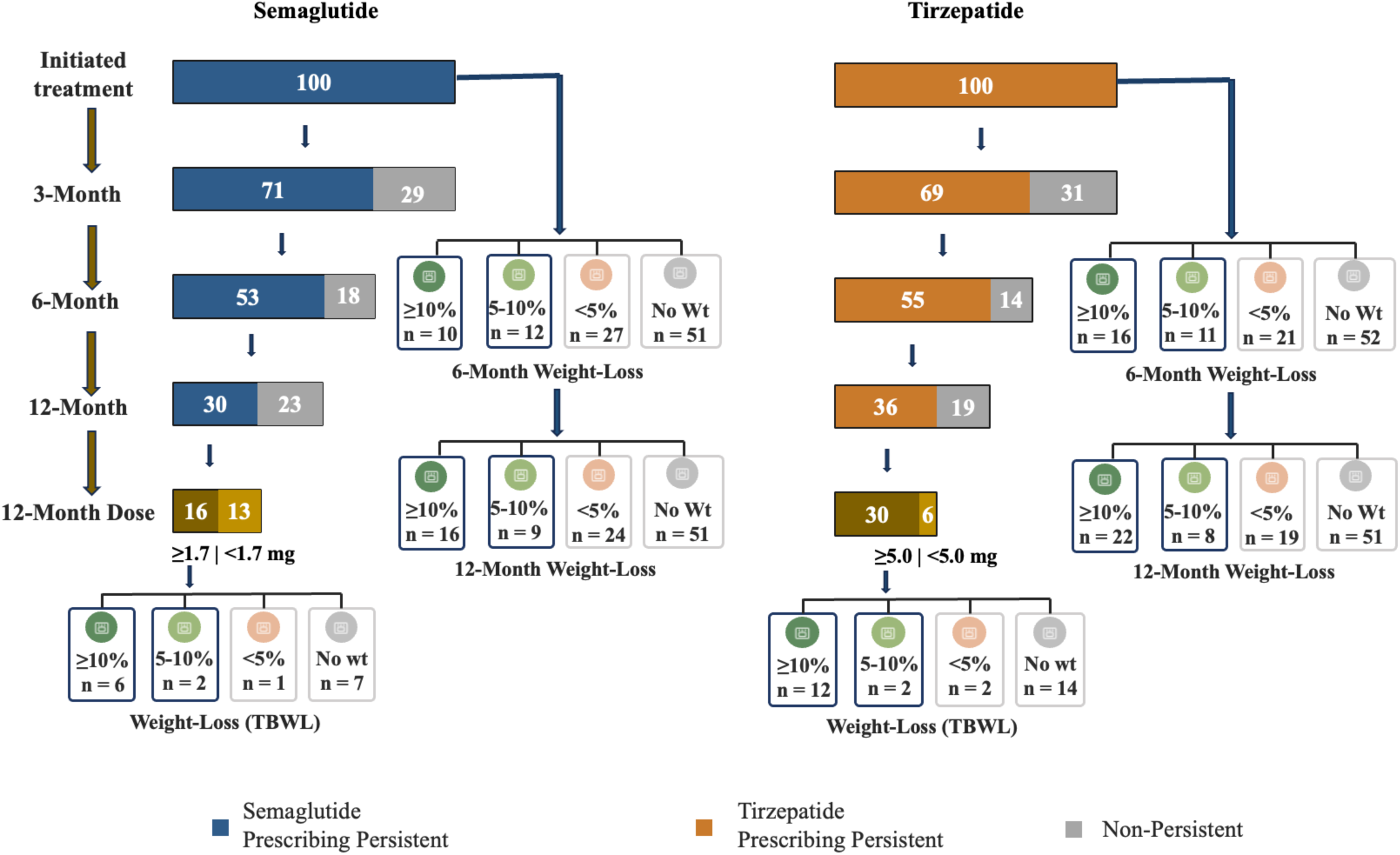

## Introduction

Semaglutide and tirzepatide have transformed the treatment of obesity. In randomized trials, adults with obesity lost on average 15% to 21% of their body weight over 68 to 104 weeks,^1–3^ and use of these medications in clinical practice has grown rapidly since their approval for chronic weight management.^4,5^ These results, however, were achieved under conditions designed to maximize benefit: participants received medication at no cost, were titrated on a fixed schedule to the highest tolerated maintenance dose, attended regular visits, and had their weight measured at each visit.

Whether patients in routine practice receive this kind of care is largely unknown. Prior real-world studies have shown that many patients discontinue these medications within the first year^6–8^ and that weight loss in clinical practice is smaller than in trials.^9^ Most of these studies, however, summarize persistence at a single time point, rely on pharmacy or insurance claims, and lack information on prescribed dose or follow-up weight. As a result, there is no detailed picture of what happens to patients after they start treatment: when they stop, whether they switch, add, or restart therapy, whether they ever reach a maintenance dose, and whether their weight is even measured. Without such a picture, it is not possible to identify where in the course of care the gap between trial efficacy and real-world effectiveness arises, or which steps: continuation, dose escalation, or monitoring, are the most important targets for improvement.

This information is needed now. Millions of US adults are starting these medications, at substantial cost to patients, health systems, and payers, and clinical guidance emphasizes structured dose escalation and regular monitoring after initiation.^10,11^ Clinicians, health systems, and policymakers making decisions about coverage and care delivery require benchmarks that describe what post-initiation care actually looks like in practice.

We therefore used electronic health record data from more than 350 US health systems to construct an atlas of the first year after initiation of semaglutide or tirzepatide among adults with obesity and without diabetes. Because prescription orders, doses, and weights are recorded in the same longitudinal record, this approach can distinguish patients who stopped treatment from those who continued but never reached a maintenance dose, and from those whose weight was never documented. For each patient, we characterized prescribing persistence, discontinuation, switching, add-on therapy, and reinitiation at 3, 6, and 12 months; attainment of FDA-labeled maintenance doses; and documentation of follow-up weight. We then examined how weight loss varied across successive steps of this care cascade and across sociodemographic groups. The study was designed to be descriptive, providing benchmarks rather than comparing the two medications.

## Methods

### Study Design and Data Source

We conducted a retrospective cohort study using the Epic Cosmos dataset, a nationwide EHR network encompassing data from more than 350 US health systems and more than 300 million patients.^12,13^ This study used de-identified data, was exempt from institutional review board review, and is reported in accordance with the Strengthening the Reporting of Observational Studies in Epidemiology (STROBE) guideline. Additional details on the data source and analytic methods are provided in the **Supplement Methods**.

### Study Population

The study population included adults aged ≥18 years who initiated injectable semaglutide or tirzepatide between January 1 and December 31, 2024 (**Supplement Figure 1**). The date of the first prescription order was defined as the index date. Individuals with prior use of either medication in any formulation were excluded. Patients were required to have at least one in-person clinical visit in the year prior to treatment initiation to ensure active engagement with the healthcare system.

The cohort was restricted to patients with obesity, defined as body mass index (BMI) ≥30 kg/m² or BMI ≥27 kg/m² with at least one obesity-related comorbidity (hypertension, dyslipidemia, or obstructive sleep apnea).^14^ We excluded individuals with a history of diabetes or prior bariatric surgery, as well as those with pregnancy or a cancer diagnosis recorded within 12 months before or after the index date. The final cohort included 446,336 semaglutide initiators and 296,817 tirzepatide initiators.

### Post-Initiation Care Patterns

Post-initiation care patterns, including prescribing persistence, discontinuation, switching, add-on/combination therapy, reinitiating, and censoring were characterized at landmark time points of 3-, 6-, and 12-months following treatment initiation. Categories were assigned hierarchically as follows: censored (death, bariatric surgery, loss of observable follow-up, or the administrative end of study follow-up on December 31, 2025), discontinued (no active weight-management medication), add-on/combination therapy (index agent plus at least one additional weight-management medication), reinitiated (renewed index agent after prior interruption), prescribing persistent (continued index agent alone without a gap exceeding 60 days), and switched (alternative agent or other anti-obesity medication active) (see **Supplement Methods** for detailed definitions). Sensitivity analyses used 30-and 90-day gaps and restricted the cohort to WEGOVY and ZEPBOUND, the Food and Drug Administration (FDA)-approved formulations for chronic weight management.

### FDA-Labeled Maintenance Dose Attainment

Initial maintenance dose attainment was defined as receipt of semaglutide ≥1.7 mg per week or tirzepatide ≥5 mg, and higher maintenance dose attainment as semaglutide ≥2.4 mg or tirzepatide ≥10 mg or ≥15 mg weekly, according to FDA labeling. We additionally described the point-in-time distribution of the prescribed weekly dose among all initiators at 30-day intervals to illustrate dose-escalation trajectories. Because prior use outside participating health systems, including compounded formulations during the 2024 drug shortages, could not be observed, some patients’ first recorded order was already at or above a maintenance dose (semaglutide, 3.8%; tirzepatide, 14.0%). These patients were counted as reaching the threshold at month 0, and a sensitivity analysis excluded them.

### Weight Monitoring and Weight Loss

Availability of follow-up weight measurements was treated as a substantive descriptive outcome rather than as a technical inclusion criterion. Total body weight loss (TBWL) was calculated as the percentage change from baseline weight to follow-up weight using the formula: TBWL (%) = (Baseline weight − Follow-up weight) / Baseline weight × 100. Weight outcomes were categorized as: ≥10% TBWL, 5-10% TBWL, <5% TBWL, and no weight recorded. TBWL was assessed both at the landmark matching each patient’s care-pattern category and at fixed 6-and 12-month landmarks across 4 progressively restricted populations: all initiators, those with recorded weight, those prescribing persistent with recorded weight, and those prescribing persistent at an FDA-labeled maintenance dose with recorded weight. For patients with multiple weight measurements within the specific window, the value closest to the target time point was used.

### Covariates

Demographic characteristics included age, sex, and race/ethnicity categorized as Hispanic, non-Hispanic (NH) White, NH Black, NH Asian, or Other. Insurance type was categorized as commercial, Medicaid or Medicaid Replacement (grouped as Medicaid), Medicare or Medicare Advantage (grouped as Medicare), self-pay, or other. Socioeconomic context was captured using the ZIP code-level Social Vulnerability Index (SVI). To characterize baseline weight trajectory, the pre-index weight change slope over the preceding 180 days was calculated.

Baseline clinical characteristics, including BMI, systolic blood pressure (SBP), and diastolic blood pressure (DBP), were ascertained during the 90-day pre-index period. Comorbid conditions were identified using International Classification of Diseases, Tenth Revision (ICD-10) codes recorded before the index date (see details in **Supplement Table 1**). Overall comorbidity burden was summarized using the age-adjusted Charlson Comorbidity Index (ACCI).^15^

## Statistical Analysis

Baseline characteristics were summarized using frequencies and percentages for categorical variables and means with standard deviations (SD) for continuous variables. Post-initiation care pattern was visualized using Sankey diagrams illustrating patient flow across mutually exclusive treatment-pattern categories at 3, 6, and 12 months, capturing attrition and treatment changes over time. Cumulative incidence of dose attainment was estimated at 0, 3, 6, and 12 months using the Aalen-Johansen estimator, with discontinuation and switching to another anti-obesity medication treated as competing events and death, bariatric surgery, loss of observable follow-up, and the administrative end of follow-up treated as censoring events. Stacked area plots showed the point-in-time distribution of the prescribed weekly dose among all initiators at 30-day intervals after initiation; unlike cumulative incidence, these proportions can decline as patients discontinue therapy. Weight-loss outcomes were reported as the proportion of patients in each predefined TBWL category. Individual-level TBWL distributions at 6 and 12 months were visualized using waterfall plots. To assess subgroup differences, multivariable logistic regression models mutually included race/ethnicity, SVI quartile, and insurance type and were additionally adjusted for states and calendar months of index date, with adjusted absolute differences estimated by marginal standardization and 95% confidence intervals (CIs) from bootstrap resampling (200 replicates). For key proportions, 95% CIs were calculated using the Wilson score method. All analyses were conducted in *R* version 4.5.1.

## Results

### Baseline Characteristics

Baseline characteristics of the 743,153 initiators [semaglutide, 446,336 (60.1%); tirzepatide, 296,817 (39.9%)] are shown in **Table 1**. Mean (SD) age was 48.5 (13.5) years; 72.5% were female, 59.5% NH White, and 68.2% commercially insured. Mean BMI was 38.8 (7.6) kg/m², with 35.7% of patients having a BMI ≥40 kg/m². Mean (SD) ACCI was 1.6 (1.8). Patients reaching later steps of the care cascade were more likely to have initiated tirzepatide and to be commercially insured; the proportion of female patients was similar across persistence steps and higher among those with recorded weight. Baseline age, BMI, and comorbidity burden were similar across care steps.

**Table 1.** Baseline Characteristics of Patients Initiating Semaglutide or Tirzepatide.

|  | **Overall\*** | **Persistent**<br>**≥3 Months** | **Persistent**<br>**≥6 Months** | **Persistent**<br>**≥12 Months** | **Reached**<br>**Maintenance**<br>**Dose at 12**<br>**Months** | **Weight**<br>**Recorded at 12**<br>**Months** | **≥5% Weight**<br>**Loss at 12**<br>**Months** |
| --- | --- | --- | --- | --- | --- | --- | --- |
| Total | 743,153 | 520,834 | 397,103 | 239,947 | 159,460 | 87,808 | 74,907 |
| Medication |  |  |  |  |  |  |  |
| Semaglutide | 446,336 (60.1%) | 314,753 (60.4%) | 234,550 (59.1%) | 133,436 (55.6%) | 70,463 (44.2%) | 39,169 (44.6%) | 32,460 (43.3%) |
| Tirzepatide | 296,817 (39.9%) | 206,081 (39.6%) | 162,553 (40.9%) | 106,511 (44.4%) | 88,997 (55.8%) | 48,639 (55.4%) | 42,447 (56.7%) |
| Age | 48.5 ± 13.5 | 48.6 ± 13.4 | 48.5 ± 13.2 | 48.4 ± 12.9 | 47.9 ± 12.5 | 48.7 ± 12.6 | 48.7 ± 12.6 |
| Sex |  |  |  |  |  |  |  |
| Female | 538,671 (72.5%) | 374,691 (71.9%) | 284,332 (71.6%) | 171,453 (71.5%) | 114,719 (71.9%) | 64,537 (73.5%) | 56,232 (75.1%) |
| Male | 204,482 (27.5%) | 146,143 (28.1%) | 112,771 (28.4%) | 68,494 (28.5%) | 44,741 (28.1%) | 23,271 (26.5%) | 18,675 (24.9%) |
| Race/Ethnicity† |  |  |  |  |  |  |  |
| Hispanic | 59,630 (8.0%) | 41,769 (8.0%) | 31,332 (7.9%) | 18,365 (7.7%) | 11,575 (7.3%) | 6,284 (7.2%) | 5,180 (6.9%) |
| NH Asian | 10,673 (1.4%) | 7,684 (1.5%) | 6,092 (1.5%) | 3,796 (1.6%) | 2,558 (1.6%) | 1,263 (1.4%) | 1,010 (1.3%) |
| NH Black | 101,992 (13.7%) | 70,113 (13.5%) | 52,882 (13.3%) | 31,377 (13.1%) | 19,094 (12.0%) | 10,806 (12.3%) | 8,573 (11.4%) |
| NH White | 442,018 (59.5%) | 310,818 (59.7%) | 237,542 (59.8%) | 144,443 (60.2%) | 98,924 (62.0%) | 55,057 (62.7%) | 47,824 (63.8%) |
| Others | 13,481 (1.8%) | 9,406 (1.8%) | 7,185 (1.8%) | 4,290 (1.8%) | 2,814 (1.8%) | 1,462 (1.7%) | 1,210 (1.6%) |
| Insurance Type |  |  |  |  |  |  |  |
| Commercial | 506,945 (68.2%) | 358,753 (68.9%) | 279,009 (70.3%) | 173,083 (72.1%) | 119,204 (74.8%) | 64,653 (73.6%) | 55,455 (74.0%) |
| Medicaid/<br>Replacement | 62,234 (8.4%) | 43,844 (8.4%) | 32,503 (8.2%) | 18,276 (7.6%) | 11,173 (7.0%) | 6,278 (7.1%) | 5,167 (6.9%) |
| Medicare/<br>Advantage | 77,285 (10.4%) | 51,450 (9.9%) | 35,130 (8.8%) | 18,588 (7.7%) | 9,477 (5.9%) | 6,198 (7.1%) | 5,191 (6.9%) |
| Self-pay | 2,772 (0.4%) | 1,978 (0.4%) | 1,499 (0.4%) | 925 (0.4%) | 466 (0.3%) | 239 (0.3%) | 189 (0.3%) |
| Other | 93,917 (12.6%) | 64,809 (12.4%) | 48,962 (12.3%) | 29,075 (12.1%) | 19,140 (12.0%) | 10,440 (11.9%) | 8,905 (11.9%) |
| SVI‡ |  |  |  |  |  |  |  |
| Quartile 1 | 152,766 (20.8%) | 109,690 (21.3%) | 86,342 (22.0%) | 54,697 (23.1%) | 38,781 (24.6%) | 20,712 (23.9%) | 18,078 (24.4%) |
| Quartile 2 | 180,573 (24.6%) | 127,622 (24.8%) | 98,298 (25.1%) | 60,093 (25.4%) | 41,127 (26.1%) | 22,540 (26.0%) | 19,503 (26.3%) |
| Quartile 3 | 185,129 (25.2%) | 128,516 (25.0%) | 97,076 (24.8%) | 58,027 (24.5%) | 38,092 (24.2%) | 21,326 (24.6%) | 18,102 (24.4%) |
| Quartile 4 | 215,689 (29.4%) | 148,616 (28.9%) | 110,435 (28.2%) | 64,127 (27.1%) | 39,556 (25.1%) | 22,239 (25.6%) | 18,377 (24.8%) |
| BMI (kg/m²) | 38.8 ± 7.6 | 38.6 ± 7.5 | 38.5 ± 7.5 | 38.4 ± 7.4 | 38.5 ± 7.3 | 38.5 ± 7.3 | 38.5 ± 7.2 |
| 27-29.9 | 41,969 (5.6%) | 30,502 (5.9%) | 23,359 (5.9%) | 14,166 (5.9%) | 8,687 (5.4%) | 4,725 (5.4%) | 3,697 (4.9%) |
| 30-34.9 | 225,027 (30.3%) | 160,177 (30.8%) | 123,481 (31.1%) | 75,526 (31.5%) | 49,891 (31.3%) | 27,007 (30.8%) | 23,040 (30.8%) |
| 35-39.9 | 210,533 (28.3%) | 147,943 (28.4%) | 113,245 (28.5%) | 68,943 (28.7%) | 46,730 (29.3%) | 25,902 (29.5%) | 22,499 (30.0%) |
| ≥40 | 265,624 (35.7%) | 182,212 (35.0%) | 137,018 (34.5%) | 81,312 (33.9%) | 54,152 (34.0%) | 30,174 (34.4%) | 25,671 (34.3%) |
| SBP | 128.0 ± 14.8 | 127.9 ± 14.7 | 127.8 ± 14.7 | 127.8 ± 14.7 | 127.7 ± 14.5 | 127.5 ± 14.4 | 127.5 ± 14.3 |
| DBP | 80.2 ± 9.6 | 80.2 ± 9.5 | 80.2 ± 9.5 | 80.3 ± 9.5 | 80.4 ± 9.5 | 80.1 ± 9.3 | 80.1 ± 9.3 |
| ACCI | 1.6 ± 1.8 | 1.6 ± 1.8 | 1.5 ± 1.7 | 1.5 ± 1.7 | 1.4 ± 1.6 | 1.5 ± 1.7 | 1.5 ± 1.7 |
| Pre-treatment weight change slope, kg/month | 0.1 ± 1.7 | 0.1 ± 1.6 | 0.1 ± 1.6 | 0.1 ± 1.6 | 0.1 ± 1.6 | 0.1 ± 1.6 | 0.1 ± 1.6 |

Plus-minus values are means ± SD. Percentages may not sum to 100 because of rounding.
\* Persistent ≥3, ≥6, and ≥12 Months indicates patients who continued the index medication alone, without a gap in prescription orders exceeding 60 days, through the corresponding landmark. Reached Maintenance Dose at 12 Months indicates patients who were prescribing persistent on the index medication at 12 months and had attained the initial FDA-labeled maintenance dose (semaglutide ≥1.7 mg or tirzepatide ≥5 mg per week). Weight Recorded at 12 Months indicates patients who met the preceding criterion and additionally had a weight measurement recorded within ±30 days of the 12-month landmark. ≥5% Weight Loss at 12 Months indicates patients who met the preceding criterion and had a total body weight loss of at least 5% from baseline.
† NH denotes non-Hispanic. Race/ethnicity was not documented for 115,359 patients (15.5% of the overall cohort).
‡ SVI denotes Social Vulnerability Index, with higher quartiles indicating greater social vulnerability. SVI was not available for 8,996 patients (1.2% of the overall cohort).
ACCI denotes the age-adjusted Charlson Comorbidity Index; BMI denotes body mass index, the weight in kilograms divided by the square of the height in meters; SBP and DBP denote systolic and diastolic blood pressure, respectively.

### Post-Initiation Care Patterns Over 12 Months

Care patterns are shown in **Figure 1 and Supplement Table 2**. Prescribing persistence declined from 70.5% (95% CI, 70.4%-70.7%) and 69.4% (95% CI, 69.3%-69.6%) at 3 months to 52.6% (95% CI, 52.4%-52.7%) and 54.8% (95% CI, 54.6%-54.9%) at 6 months, and to 29.9% (95% CI, 29.8%-30.0%) and 35.9% (95% CI, 35.7%-36.1%) at 12 months, for semaglutide and tirzepatide respectively; discontinuation and switching accounted for most attrition. Combined prescribing persistence of semaglutide and tirzepatide declined from 70.1% (95% CI, 70.0%-70.2%) at 3 months to 32.3% (95% CI, 32.2%-32.4%) at 12 months (**Supplement Figure 2**), with similar results using 30-and 90-day gap definitions (**Supplement Figures 3-4**). Twelve-month prescribing persistence was 25.8% (95% CI, 25.6%-26.0%) among WEGOVY initiators and 31.2% (95% CI, 31.0%-31.4%) among ZEPBOUND initiators, lower than among all injectable semaglutide and tirzepatide initiators (**Supplement Figure 5, Supplement Table 3**).

**Figure 1.**
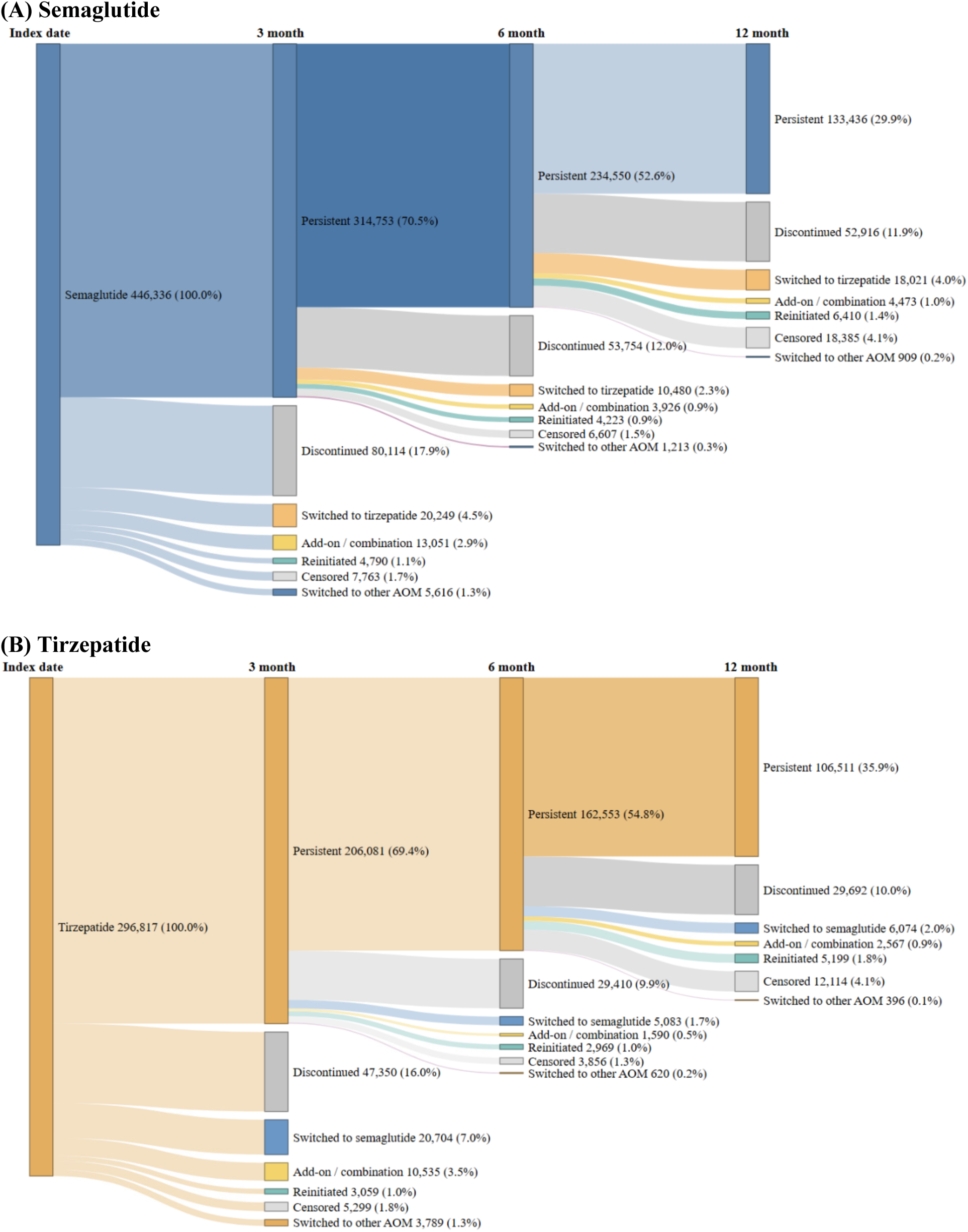
Real-World Care Patterns after Initiating Semaglutide or Tirzepatide for Obesity. Shown are Sankey diagrams depicting the flow of patients across post-initiation care categories at the index date and at 3, 6, and 12 months after initiating (A) semaglutide or (B) tirzepatide. Once a patient exited the persistent category at a given landmark, that patient’s subsequent status was not re-classified at later landmarks; band widths are proportional to the number of patients in each category.

Among all initiators, prescribing persistence at 12 months was 29.9% for semaglutide and 35.9% for tirzepatide; this fell to 15.8% (95% CI, 15.7%-15.9%) and 30.0% (95% CI, 29.8%-30.1%) among those additionally reaching maintenance dose, and to 8.8% (95% CI, 8.7%-8.9%) and 16.4% (95% CI, 16.3%-16.5%) among those further having a weight recorded within the 12-month assessment window, reflecting substantial attrition earlier in the cascade of care (**Supplement Figures 6-7**).

Prescribing persistence at 12 months varied across subgroups (**Supplement Figures 6-7**). By race/ethnicity, prescribing persistence was highest among NH Asian patients (32.2% for semaglutide, 40.6% for tirzepatide) and lowest among Hispanic semaglutide initiators (29.4%) and NH Black tirzepatide initiators (32.2%). By SVI quartile, prescribing persistence was highest in quartile 1 (32.2% and 40.5%) and lowest in quartile 4 (28.4% and 32.1%). By insurance type, prescribing persistence was highest among self-pay semaglutide initiators (33.4%) and commercially insured tirzepatide initiators (37.6%), and lowest among Medicare-insured patients for both medications (23.2% and 25.9%). Similar patterns with slightly higher attrition were observed among WEGOVY and ZEPBOUND initiators (**Supplement Figures 8-9**).

### FDA-Labeled Maintenance Dose Attainment and Dose Escalation

Cumulative FDA-labeled maintenance dose attainment among all initiators, irrespective of persistence status, is shown in **Figure 2**. On the index date, 3.8% of semaglutide and 14.0% of tirzepatide initiators already had an order at or above the initial maintenance dose. Among semaglutide initiators, 28.3% (95% CI, 28.2%-28.4%) ever reached ≥1.7 mg and 16.9% (95% CI, 16.8%-17.0%) ≥2.4 mg by 12 months.

**Figure 2.**
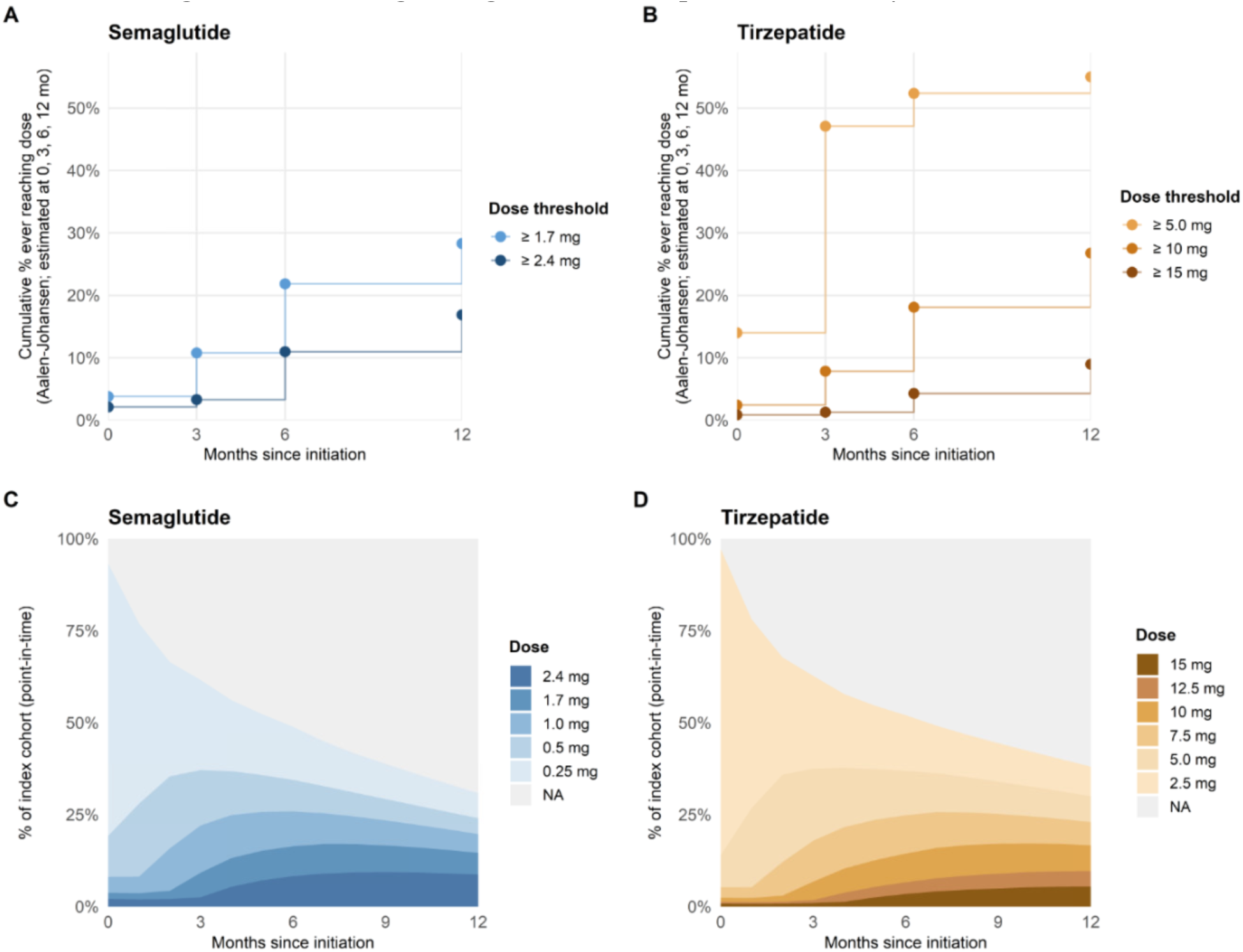
FDA-Labeled Maintenance-Dose Attainment and Monthly Dose Distribution Over 12 Months Among Adults Initiating Semaglutide or Tirzepatide for Obesity. (A, B) Cumulative incidence of ever reaching each weekly dose threshold for semaglutide (≥1.7 mg, the initial FDA-labeled maintenance dose, and ≥2.4 mg) and tirzepatide (≥5 mg, the initial FDA-labeled maintenance dose, and ≥10 mg and ≥15 mg), estimated with the Aalen-Johansen estimator at 0, 3, 6, and 12 months. Discontinuation and switching to another anti-obesity medication were treated as competing events; censoring was reserved for death, bariatric surgery, loss of observable follow-up, and administrative end of study follow-up. (C, D) Point-in-time distribution of the prescribed weekly dose among all initiators at monthly intervals after initiation. *NA* indicates no active order (including after discontinuation) or missing dose. FDA indicates Food and Drug Administration.

Among tirzepatide initiators, 55.0% (95% CI, 54.8%-55.2%) reached ≥5 mg, 26.8% (95% CI, 26.6%-26.9%) ≥10 mg, and 9.0% (95% CI, 8.9%-9.1%) ≥15 mg by 12 months. Among patients prescribing persistent at 12 months, maintenance attainment was substantially higher (semaglutide 1.7 mg: 52.8% [95% CI, 52.5%-53.1%]; tirzepatide 5.0 mg: 83.6% [95% CI, 83.3%-83.8%]) (**Supplement Figure 10**). In a sensitivity analysis excluding patients whose first recorded order was already at or above the initial maintenance dose (semaglutide, n = 17,015 [3.8%]; tirzepatide, n = 41,593 [14.0%]), 12-month attainment was 25.5% (95% CI, 25.3%-25.6%) for semaglutide and 47.7% (95% CI, 47.5%-47.9%) for tirzepatide. Among all initiators, 12-month attainment of the initial maintenance dose was highest among NH Asian patients (semaglutide, 32.0%; tirzepatide, 60.4%) and those in SVI quartile 1 (31.4% and 59.9%), and lowest among Medicare-insured patients (16.5% and 39.4%) (**Supplement Figure 11**). A similar pattern was observed for the higher labeled maintenance dose (semaglutide, ≥2.4 mg; tirzepatide, ≥15 mg), with the lowest attainment among Medicare-insured patients and, for semaglutide, self-pay patients (**Supplement Figure 12**).

### Weight-Loss Response According to Completeness of Care

Weight-loss outcomes by care pattern are shown in **Figures 3A-3B** and **Supplement Tables 4-5**. About half of initiators lacked a recorded weight at each landmark (range, 44.4%-54.1%). Response rose consistently with the completeness of post-initiation care. Among patients who were not prescribing persistent, ≥5% TBWL was recorded in 10.7% (95% CI, 10.5%-10.8%), 15.5% (95% CI, 15.2%-15.7%), and 22.9% (95% CI, 22.6%-23.2%) of semaglutide initiators evaluated at the 3-, 6-, and 12-month non-persistence landmarks, respectively, and in 11.5% (95% CI, 11.3%-11.8%), 17.3% (95% CI, 16.9%-17.7%), and 26.4% (95% CI, 26.0%-26.7%) of tirzepatide initiators. Among patients who remained prescribing persistent through 12 months, response was substantially greater, and further differed by dose attainment: those who had reached the initial FDA-labeled maintenance dose recorded ≥5% TBWL approximately twice as often or more than those who had not (semaglutide, 46.1% [95% CI, 45.7%-46.4%] vs 23.4% [95% CI, 23.1%-23.8%]; tirzepatide, 47.7% [95% CI, 47.4%-48.0%] vs 17.1% [95% CI, 16.6%-17.7%]), similar for ≥10% TBWL (semaglutide, 34.9% [95% CI, 34.5%-35.2%] vs 13.7% [95% CI, 13.4%-13.9%]; tirzepatide, 40.4% [95% CI, 40.0%-40.7%] vs 10.0% [95% CI, 9.6%-10.5%]).

**Figure 3A.**
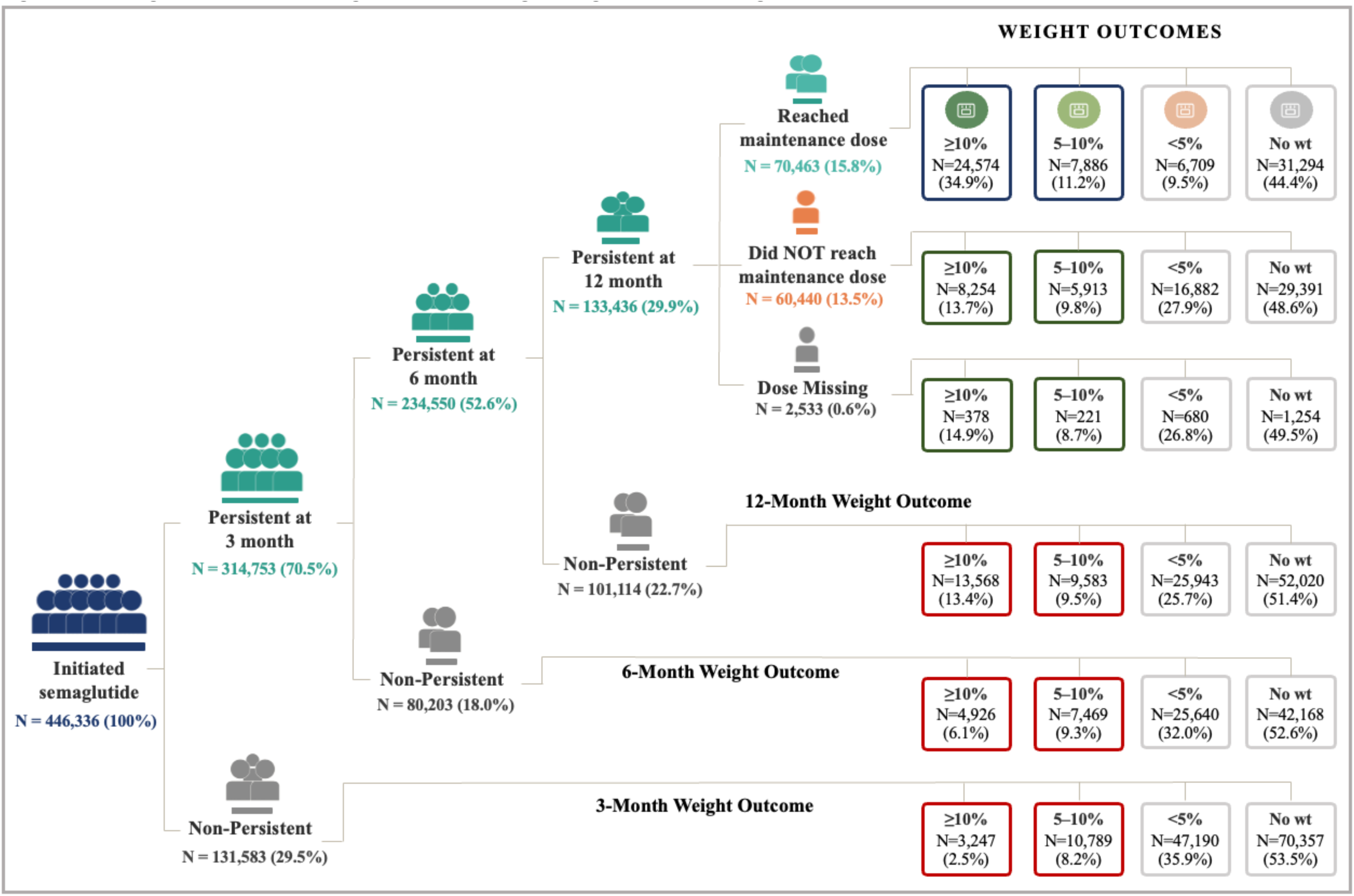
Weight Outcomes Among Adults Initiating Semaglutide According to Post-Initiation Care Patterns.

**Figure 3B.**
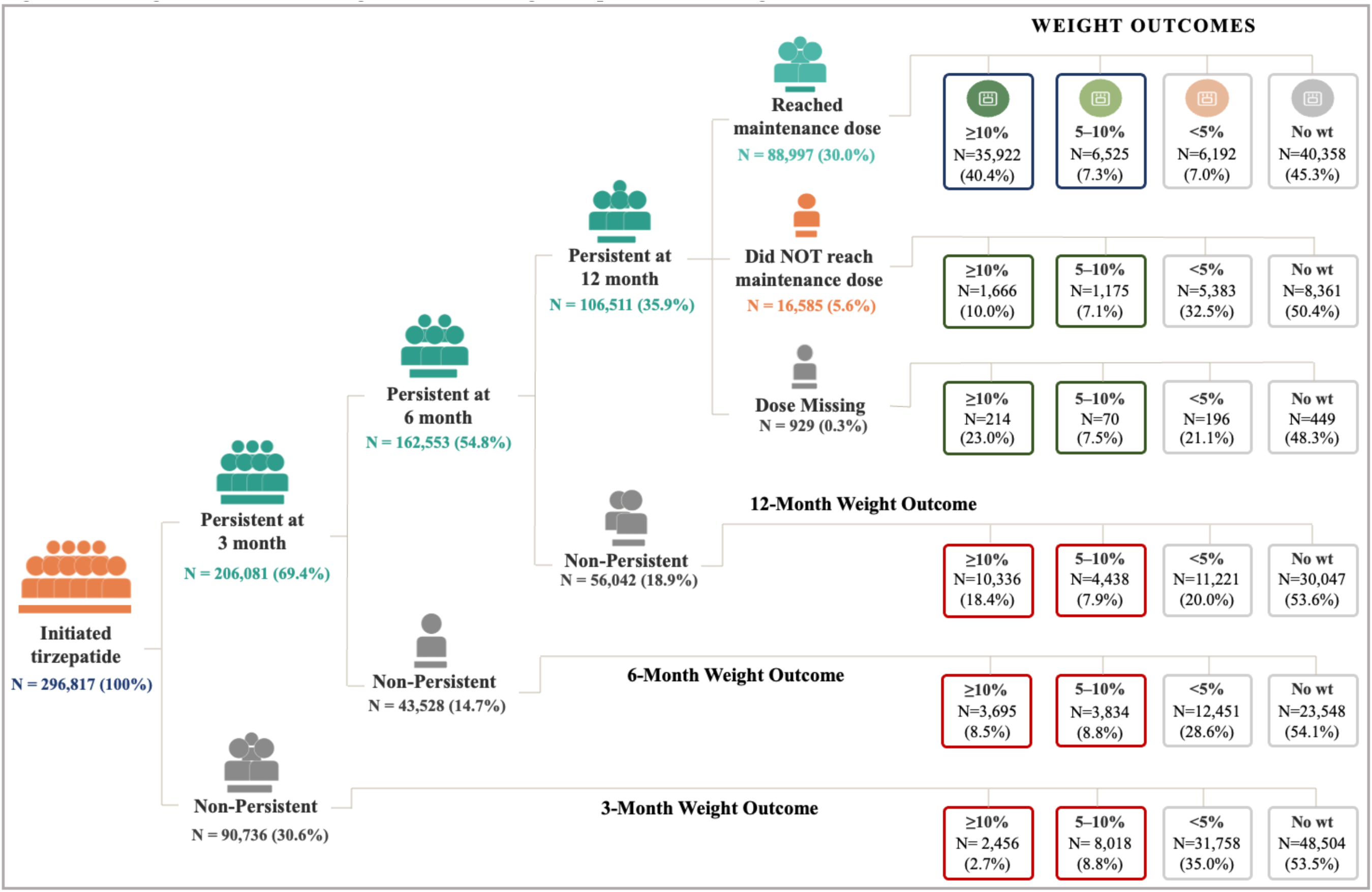
Weight Outcomes Among Adults Initiating Tirzepatide According to Post-Initiation Care Patterns. Shown is the distribution of total body weight loss (TBWL) at 3, 6, and 12 months among (3A) semaglutide initiators and (3B) tirzepatide initiators, stratified by post-initiation care pattern. Weight outcomes were categorized as at least 10% TBWL, 5 to 10% TBWL, less than 5% TBWL, or no weight recorded.

### Weight Loss Outcomes at Fixed 6-and 12-Month Landmarks

The proportion of patients with recorded ≥5% TBWL rose progressively across four increasingly restricted, nested populations: all initiators, those with a recorded weight, those also prescribing persistent, and those further at a maintenance dose, at both 6 and 12 months (**Figure 4; Supplement Figure 13; Supplement Tables 6-7**). Among all initiators, including those without a recorded weight, ≥5% TBWL at 12 months was recorded in 25.6% (95% CI, 25.5%-25.7%) of semaglutide and 30.4% (95% CI, 30.2%-30.6%) of tirzepatide users. This rose to 51.9% (95% CI, 51.7%-52.1%) and 61.9% (95% CI, 61.6%-62.1%) among those with a recorded weight, to 66.1% (95% CI, 65.7%-66.4%) and 79.5% (95% CI, 79.1%-79.8%) among those additionally prescribing persistent, and to 82.9% (95% CI, 82.5%-83.2%) and 87.3% (95% CI, 87.0%-87.6%) among those further at a labeled maintenance dose, with similar trends for ≥10% TBWL and at 6 months.

**Figure 4.**
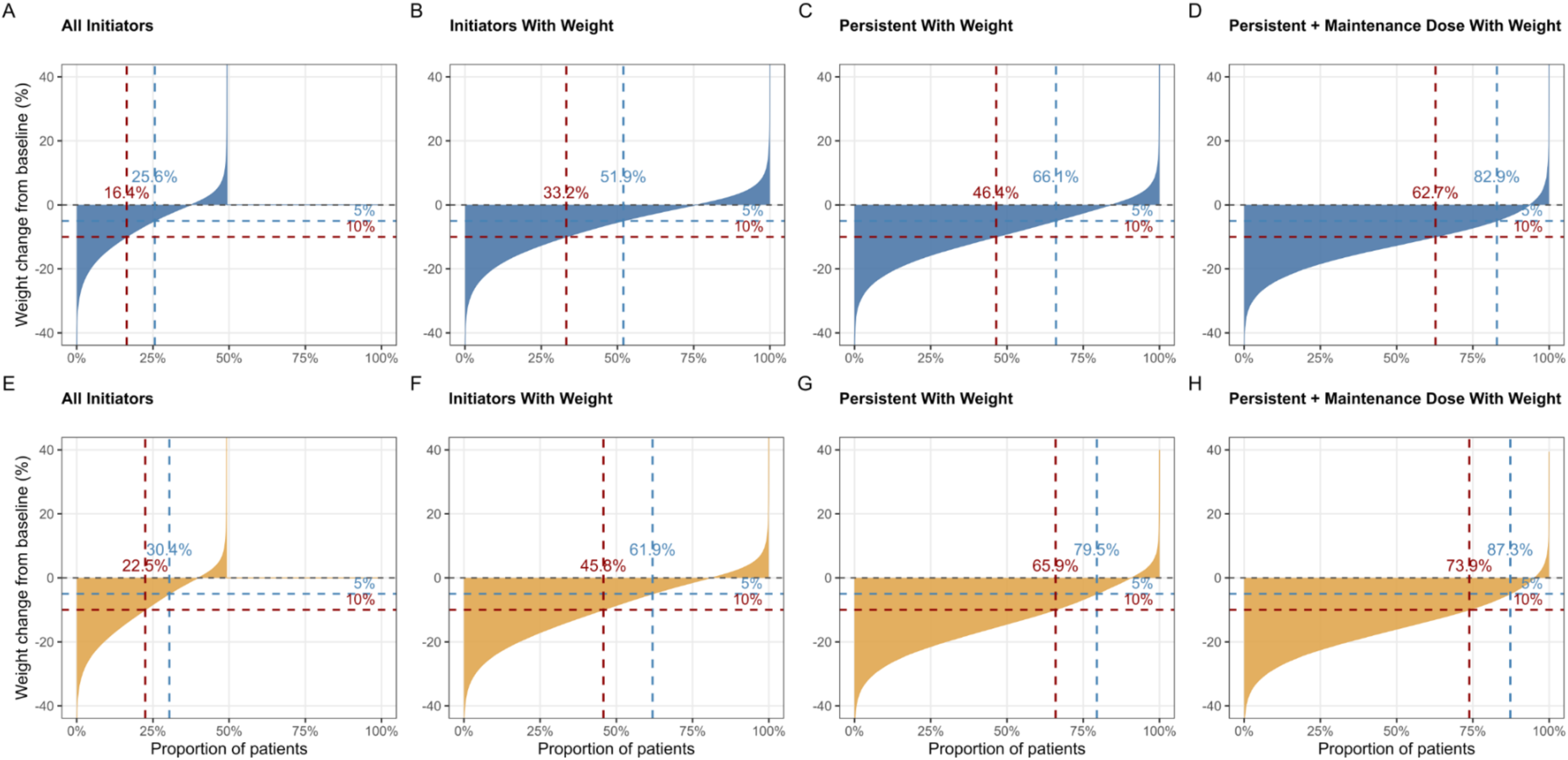
Weight Change Among Adults Initiating Semaglutide or Tirzepatide for Obesity At 12 Months. Shown is the distribution of percent total body weight change from baseline at 12 months among (A, E) all initiators, (B, F) initiators with a recorded weight, (C, G) initiators who were prescribing persistent with a recorded weight, and (D, H) initiators who were prescribing persistent at an FDA-labeled maintenance dose with a recorded weight, for (A-D) semaglutide and (E-H) tirzepatide. Each curve represents the distribution of percent weight change from baseline, with patients ordered from greatest to least weight loss; percentages shown indicate the proportion of patients achieving at least 5% (blue) or 10% (red) weight loss.

Lower response was generally observed among male patients, NH Black patients, and those in higher-SVI areas across care-pattern groups. Differences by insurance type were less consistent; for example, among all initiators, Medicare beneficiaries more often had ≥5% TBWL documented than commercially insured patients, reflecting more frequent weight documentation. When the analysis was restricted to WEGOVY and ZEPBOUND initiators, the same care-cascade gradient and subgroup disparities were evident, with response only modestly attenuated relative to all initiators (**Supplement Tables 8-9**).

### Adjusted Differences by Race, Insurance, and Social Vulnerability

As shown in **Supplement Table 10**, after multivariable adjustment, absolute differences (in percentage points) by race/ethnicity were modest for prescribing persistence and dose attainment (within 2 points for semaglutide, about 3 for tirzepatide) but persisted for weight response (NH Black vs NH White, semaglutide/tirzepatide:-8.8 [95% CI,-9.4 to-8.0] and-9.2 [95% CI,-10.2 to-8.2]). A graded association with SVI quartile persisted across all three outcomes (for example, Q4 vs Q1, semaglutide/tirzepatide: prescribing persistence,-3.7 [-4.2 to-3.2] and-6.6 [-7.2 to-6.0]). Differences by insurance type were largest for Medicare (for example, semaglutide/tirzepatide: prescribing persistence, - 8.4 [-8.8 to-7.9] and-12.1 [-12.6 to-11.5]).

## Discussion

In this nationwide cohort of more than 740,000 adults initiating semaglutide or tirzepatide for obesity, we found substantial attrition across successive steps of post-initiation care. Only about one-third of patients remained prescribing persistent at 12 months, approximately half had a documented follow-up weight measurement, and only about one-quarter of all initiators observed clinically meaningful weight loss. Weight loss increased progressively across more complete care trajectories, among patients who remained prescribing persistent at an FDA-labeled maintenance dose and had recorded follow-up weight, more than 80% achieved at least 5% weight loss. These findings indicate that post-initiation care of semaglutide or tirzepatide therapy in routine practice is marked by substantial gaps in prescribing persistence, maintenance-dose attainment, and weight follow-up, and that the real-world effectiveness of these therapies may depend in part on successful delivery of that care. However, causal interpretation warrants caution given the potential for selection bias, and future studies using designs suited to time-varying are needed.

Our findings help explain the gap between the efficacy reported in clinical trials and the effectiveness observed in routine practice. In the STEP and SURMOUNT programs, mean weight reductions of 15%-21% were achieved under conditions including reliable medication access, structured dose escalation, behavioral support, and systematic outcome assessment.^1–3,16^ In routine care, patients face barriers including high out-of-pocket costs, limited insurance coverage, restricted access to follow-up care, and even medication shortages.^6–8^ The observed disparities across social vulnerability, race/ethnicity, and insurance type are also consistent with the hypothesis that barriers to longitudinal care contribute to lower real-world treatment effectiveness. Higher social vulnerability was generally associated with lower prescribing persistence, maintenance-dose attainment, and documented weight-loss response; NH Black patients had lower maintenance-dose attainment and weight-loss response than NH White patients, while differences in prescribing persistence varied by medication and patterns by insurance type varied across outcomes.^13^ These findings identify populations that may benefit from interventions designed to improve continuity of treatment and follow-up.

These findings suggest that many patients discontinue therapy without transitioning to an alternative evidence-based treatment.^17–20^ Because the study period overlapped with widespread availability of compounded GLP-1 formulations through telehealth platforms and online pharmacies, some patients classified as having discontinued treatment may have transitioned to compounded products obtained outside participating health systems, leading to an overestimation of treatment discontinuation in the electronic health record.^21^ This may also partly explain why some patients classified as nonpersistent at the 3-or 6-month landmark nonetheless achieved clinically meaningful weight loss.

One notable finding was the high proportion of patients without documented follow-up weight measurements. Although some patients may have been monitored outside participating health systems or through home measurements not captured in the electronic health record, the absence of documented weight limits assessment of treatment response and may itself represent an important gap in routine obesity care. Regular assessment of weight is fundamental to evaluating treatment effectiveness and guiding decisions regarding dose escalation, continuation, or alternative therapies.

These findings have several implications. Supporting persistence, successful dose escalation, and structured follow-up may be important for improving real-world treatment outcomes. However, these findings should not be interpreted as demonstrating a causal effect of post-initiation care on weight-loss outcomes. Patients who remained persistent on therapy, reached maintenance doses, and returned for follow-up weight assessments likely represent a selected subgroup with better treatment tolerance, greater engagement with care, or more favorable early treatment response.^8,22^ Conversely, patients with limited weight loss, poor tolerability, or barriers to care may have been more likely to discontinue treatment or miss follow-up visits. Therefore, the observed associations may reflect both the effects of longitudinal care and underlying differences in patient characteristics. Future studies using causal inference approaches are needed to disentangle these mechanisms. Furthermore, the substantial proportion of patients without documented follow-up weight also highlights the need for more systematic outcome monitoring in routine obesity care. Without objective assessment of treatment response, clinicians cannot make informed decisions regarding treatment continuation, dose escalation, or switching therapies. Finally, tirzepatide consistently demonstrated higher weight-loss response rates than semaglutide among all initiators and among persistent users. However, these comparisons should be interpreted cautiously because the recommended maintenance-dose range for tirzepatide is substantially broader than that for semaglutide, and differences in dose escalation may partially account for the observed differences in treatment response.

This study has several limitations. Medication exposure was based on prescription orders rather than confirmed dispensing or medication use, therefore, persistence reflects continuity of prescribing rather than adherence. Weight measurements and medication use occurring outside participating health systems were not captured, and some patients may have obtained compounded GLP-1 products or received follow-up care outside the electronic health record network. The study also lacked information on reasons for treatment discontinuation, such as adverse effects, lifestyle interventions, medication costs, or insurance coverage decisions. Finally, follow-up was limited to 12 months and does not capture longer-term treatment patterns or outcomes.

In conclusion, substantial attrition occurred across successive steps of post-initiation care for semaglutide or tirzepatide based obesity therapies in routine clinical practice, including prescribing persistence, dose escalation, and weight monitoring. These findings provide insights into potential contributors to the gap between clinical trial efficacy and real-world effectiveness. Reliable medication access, structured follow-up, appropriate dose escalation, and routine outcome measurement represent opportunities to close these observed gaps and reduce inequities in access to obesity pharmacotherapy.

## Data Sharing Statement

Data are proprietary to Epic Cosmos and are not publicly available. Access requests can be directed to the Epic Cosmos Research Network.

