## Supplementary Appendix for "Post-Initiation Care Patterns and Weight Loss With Semaglutide and Tirzepatide for Obesity in Clinical Practice"

### Supplement Methods

### Supplement References

### Supplement Results

**Supplement Figure 1.** Flow Chart of Participants.

**Supplement Figure 2.** Post-Initiation Care Pattern from Treatment Initiation To 12 Months Among Patients Initiating Semaglutide or Tirzepatide.

**Supplement Figure 3.** Sensitivity Analysis of Post-Initiation Care Pattern Using A 30-Day Gap Between Prescription Orders.

**Supplement Figure 4.** Sensitivity Analysis of Post-Initiation Care Pattern Using A 90-Day Gap Between Prescription Orders.

**Supplement Figure 5.** Sensitivity Analysis of Post-Initiation Care Pattern Among Patients Initiating WEGOVY or ZEPBOUND.

**Supplement Figure 6.** Stepwise Attrition Along the Post-Initiation Care Continuum Among Semaglutide Initiators, Overall and by Race/Ethnicity, Social Vulnerability Index Quartile, and Insurance Type.

**Supplement Figure 7.** Stepwise Attrition Along the Post-Initiation Care Continuum Among Tirzepatide Initiators, Overall and by Race/Ethnicity, Social Vulnerability Index Quartile, and Insurance Type.

**Supplement Figure 8.** Stepwise Attrition Along the Post-Initiation Care Continuum Among WEGOVY Initiators, Overall and by Race/Ethnicity, Social Vulnerability Index Quartile, and Insurance Type.

**Supplement Figure 9.** Stepwise Attrition Along the Post-Initiation Care Continuum Among ZEPBOUND Initiators, Overall and by Race/Ethnicity, Social Vulnerability Index Quartile, and Insurance Type.

**Supplement Figure 10.** FDA-Labeled Maintenance-Dose Attainment and Monthly Dose Distribution Over 12 Months Among Initiators Prescribing Persistent Through 12 Months.

**Supplement Figure 11.** Cumulative Incidence of Reaching the Initial FDA-Labeled Maintenance Dose (Semaglutide: 1.7 mg; Tirzepatide: 5 mg) by Subgroups.

**Supplement Figure 12.** Cumulative Incidence of Reaching the Higher FDA-Labeled Maintenance Dose (Semaglutide: 2.4 mg; Tirzepatide: 15 mg) by Subgroups.

**Supplement Figure 13.** Weight Change Among Adults Initiating Semaglutide or Tirzepatide for Obesity At 6 Months.

**Supplement Table 1.** Definitions of Study Variables and Corresponding Codes.

**Supplement Table 2.** Post-Initiation Care Over 12 Months With 95% Confidence Intervals.

**Supplement Table 3.** Sensitivity Analyses of Post-Initiation Care Over 12 Months With 95% Confidence Intervals.

**Supplement Table 4.** Weight-Loss Outcomes of Semaglutide Initiators by Care Pattern.

**Supplement Table 5.** Weight-Loss Outcomes of Tirzepatide Initiators by Care Pattern.

**Supplement Table 6.** Weight-Loss Outcomes at 6- and 12-Month of Semaglutide Initiators.

**Supplement Table 7.** Weight-Loss Outcomes at 6- and 12-Month of Tirzepatide Initiators.

**Supplement Table 8.** Weight-Loss Outcomes at 6- and 12-Month of WEGOVY Initiators.

**Supplement Table 9.** Weight-Loss Outcomes at 6- and 12-Month of ZEPBOUND Initiators.

**Supplement Table 10.** Adjusted Absolute Differences (in Percentage Points) in Prescribing Persistence, Maintenance Dose Attainment, and Weight Loss by Race and Ethnicity, Social Vulnerability Index, and Insurance Type.

### Supplement Methods

#### Data Source

This study used Epic Cosmos, a deidentified electronic health record (EHR) research network that aggregates data from more than 350 contributing US health systems on the Epic platform, together representing an aggregate population of more than 300 million patients across all 50 states.[1] A shared patient-linkage identifier is applied across contributing organizations, so that encounters, diagnoses, and orders recorded for the same individual at different participating systems are consolidated into a single longitudinal record rather than treated as separate episodes of care. Care delivered exclusively outside participating systems is not captured, and the roster of contributing organizations has continued to expand since the network's inception.

**Figure. Proportions of patients represented in Cosmos compare to the United States**

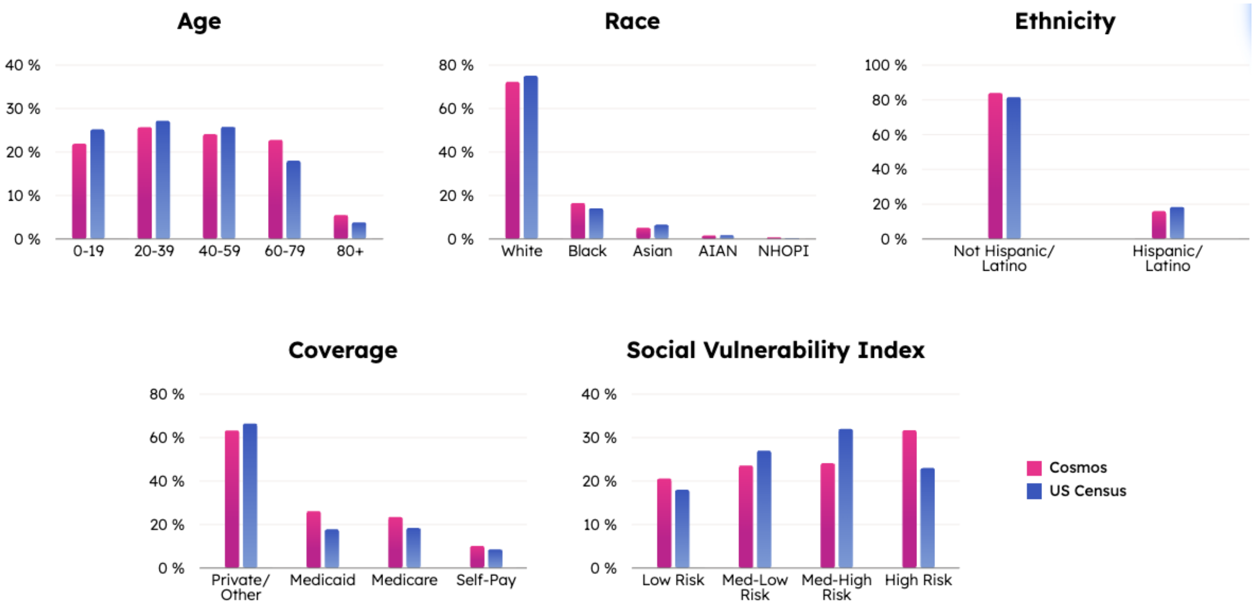


(Epic Cosmos webpage, https://cosmos.epic.com/about/)

Data released for research use are deidentified under the HIPAA Safe Harbor provision, with direct identifiers removed before release. To limit re-identification risk while preserving the ability to measure elapsed time between clinical events, every date in a given patient's record is shifted by a single random offset (0-30 days) unique to that patient; because the same offset is applied uniformly across all of that patient's records, the interval between any two events for that individual (eg, an index prescription order and a subsequent encounter) is unaffected even though the underlying calendar date cannot be recovered.

**Cohort Definitions**

Obesity status was determined from the qualifying pre-index body mass index (BMI) using a threshold of BMI 30 kg/m² or greater, or 27 kg/m² or greater with at least one obesity-related comorbid condition (hypertension, dyslipidemia, or obstructive sleep apnea).

Diabetes before the index date was ascertained using either an International Classification of Diseases, Tenth Revision (ICD-10) diagnosis code in the E08-E11 or E13 range or a prescription order for an anti-diabetic medication. The anti-diabetic medication list used for this purpose excluded injectable semaglutide and tirzepatide themselves. Patients were also excluded for a history of bariatric surgery, for pregnancy recorded within 12 months before or after the index date, or for a cancer diagnosis within that same window, since each of these conditions can independently affect body weight through mechanisms unrelated to GLP-1 receptor agonist or GIP/GLP-1 receptor co-agonist pharmacotherapy.

The index date was defined as the date of a patient's first injectable semaglutide or tirzepatide prescription order in calendar year 2024. To restrict the cohort to treatment-naive initiators, patients with any earlier order for either medication in any formulation, including oral semaglutide, were excluded. Patients were additionally required to have at least one in-person encounter in the 12 months preceding the index date, so that all included patients had a documented point of contact with a participating health system before treatment initiation.

**Ascertainment of Vital Signs and Medications**

Weight, height, BMI, and blood pressure were obtained from structured vital-sign flowsheet entries generated during any encounter type, including office visits, telehealth, and laboratory or procedural visits. Physiologically implausible entries, weight below 20 kg or above 300 kg, height below 0.5 m or above 3 m, or BMI below 15 kg/m² or above 120 kg/m², were excluded before any downstream calculation. When multiple qualifying measurements fell on the same calendar day, they were averaged into a single daily value; when multiple qualifying daily values fell within a given assessment window, the value closest to the window's target date was retained.

Exposure to semaglutide and tirzepatide and to other anti-obesity medications was identified from structured prescription order entries, which record generic and product name, ordered dose, dose unit, and route. Because Cosmos captures what was prescribed rather than what was filled or actually taken, a prescription order indicates clinical intent rather than confirmed receipt or use of the medication. Weekly dose was parsed directly from the structured dose field on each order; orders without a structured dose value were coded as having missing dose information. Dose was recorded in milligrams for 92.4% of semaglutide and 97.0% of tirzepatide index orders; values recorded in other units were treated as missing.

**Classification of Post-Initiation Care Pattern**

Care pattern was reassessed at three landmarks after the index date (3, 6, and 12 months) each evaluated within a ±30-day window. At every landmark, patients were assigned to a treatment-pattern category using a fixed hierarchy of rules evaluated in sequence, such that a patient satisfying an earlier rule was not evaluated against subsequent rules: (1) censored, for death, bariatric surgery, absence of any further recorded encounter through the end of the observation period, or arrival at the administrative data cutoff of December 31, 2025; (2) discontinued, for patients with no active order for the index medication, or any other anti-obesity medication during the landmark window; (3) add-on/combination therapy, for an active index-medication order overlapping with an order for at least one additional anti-obesity medication; (4) reinitiated, for resumption of index-medication-alone prescribing after a gap exceeding 60 days; (5) prescribing persistent, for continued index-medication-alone prescribing, with no gap exceeding 60 days; and (6) switched, for an active order for semaglutide, tirzepatide, or another anti-obesity medication in the absence of any active index-medication order. Because re-evaluation at each landmark was restricted to patients persistent at the prior landmark, reinitiated captured only a gap-and-restart cycle within a single inter-landmark interval, not later resumption by patients already coded discontinued or switched at an earlier landmark, who remained fixed in that category by design. Reported category proportions at 6 and 12 months describe the subsequent trajectory of the subset of patients who remained prescribing persistent at the immediately preceding landmark, not an independent re-evaluation of the full baseline cohort at each time point.

The 60-day allowable gap served as the primary prescribing persistence definition, 30-day and 90-day gap thresholds were applied in sensitivity analyses. An additional sensitivity analysis restricted the cohort to initiators of WEGOVY or ZEPBOUND, the formulations of semaglutide and tirzepatide, respectively, that carry an FDA indication specifically for chronic weight management, to evaluate whether findings were consistent when non-labeled formulations were excluded.

**FDA-Labeled Maintenance Dose Attainment**

An initial FDA-labeled maintenance dose was defined as a prescribed weekly dose of at least 1.7 mg for semaglutide or at least 5.0 mg for tirzepatide, and a higher labeled maintenance dose as at least 2.4 mg for semaglutide or at least 10 mg and 15 mg for tirzepatide. Cumulative attainment of each threshold was estimated using the Aalen-Johansen estimator, with discontinuation or switching before dose attainment treated as a competing event and death, bariatric surgery, loss of observable follow-up, and reaching the administrative data cutoff treated as censoring events. FDA-labeled dose attainment reflects at least one qualifying order at or above the threshold, not sustained use. For each landmark, the highest prescribed weekly dose was mapped to the nearest labeled dose step (semaglutide 0.25, 0.5, 1.0, 1.7, 2.0, and 2.4 mg; tirzepatide 2.5 to 15 mg in 2.5-mg steps) when within 8% of that step, and other values were treated as missing; orders for semaglutide 2.0 mg therefore counted toward the ≥1.7 mg threshold. Patients with an order at or above a threshold on the index date were counted as reaching it at month 0; because no competing events or censoring can occur before the index date, the month-0 estimate equals the crude proportion. Monthly dose distributions were constructed at 30-day intervals from the index date using the highest dose among orders active on that day; patients without an active order were shown as having no active order or missing dose. Notably, attainment does not imply clinical optimality: because titration is individualized to tolerability and physician judgment, patients appropriately maintained below a labeled maintenance dose are still coded as not having reached one.

**Ascertainment of Weight Outcomes**

Availability of a follow-up weight measurement was analyzed as an outcome in its own right rather than treated solely as an analytic prerequisite, because incomplete weight ascertainment can reflect disengagement from care and, if not distributed at random across subgroups, can bias real-world estimates of treatment effectiveness derived only from patients with recorded follow-up weight. A patient was considered to have a qualifying follow-up weight at a given landmark if any structured weight measurement fell within ±30 days of that landmark, patients without such a measurement were reported as missing.

Among patients with a qualifying follow-up weight, percent total body weight loss (TBWL) was calculated as (baseline weight − follow-up weight) / baseline weight × 100, with baseline weight defined as the value closest to the index date within the preceding 90 days. TBWL was examined both as a continuous measure and in 3 prespecified categories: 10% or greater loss, 5% to less than 10% loss, and less than 5% loss (including any weight gain), with a threshold of 5% or greater serving as the primary definition of clinically meaningful weight-loss response and a threshold of 10% or greater examined as a secondary, more stringent definition. 95% confidence intervals for key proportions were calculated using the Wilson score method.

Weight-loss outcomes were examined in two complementary ways. First, within the post-initiation care pattern classification, each patient's TBWL was evaluated at the single landmark corresponding to that patient's observed care-pattern category, allowing weight-loss distributions to be compared directly across mutually exclusive care-pattern subgroups. Second, TBWL was assessed at the fixed 6- and 12-month landmarks across 4 progressively restricted populations: (1) all initiators, regardless of follow-up weight availability; (2) initiators with a recorded follow-up weight; (3) initiators prescribing persistent with the index medication who also had a recorded follow-up weight; and (4) initiators prescribing persistent with the index medication at an FDA-labeled maintenance dose who also had a recorded follow-up weight.

**Sociodemographic and Clinical Covariates**

Race and ethnicity were derived from the structured Race and Ethnicity fields recorded in the patient record. Patients recorded as Hispanic or Latino on the Ethnicity field were classified as Hispanic irrespective of the accompanying Race value; remaining patients were classified as non-Hispanic White, non-Hispanic Black, non-Hispanic Asian, or Other according to their recorded Race field, and patients with neither field documented were retained as a separate undocumented category rather than being excluded or imputed.

Insurance type reflected the primary payer on file for the encounter associated with the index prescription order and was grouped as commercial, Medicaid or Medicaid Replacement (grouped as Medicaid), Medicare or Medicare Advantage (grouped as Medicare), self-pay, or other.

Neighborhood-level social disadvantage was captured using ZIP code-level Social Vulnerability Index (SVI) values derived from the 2020 SVI release published by the Centers for Disease Control and Prevention, linked to each patient through the ZIP code recorded as their residence and appended within the Cosmos data infrastructure.

Pre-index weight trajectory was summarized as a linear slope (kg per month) fit by ordinary least-squares regression of weight on time-to-index, using all qualifying weight measurements recorded from 180 days before the index date; a positive slope denotes pre-index weight gain and a negative slope denotes pre-index weight loss. Patients with fewer than 2 qualifying pre-index weights, or whose available measurements spanned no variation in time from the index date, could not support a slope estimate and were coded as missing for this covariate. To limit the influence of extreme or implausible slope estimates, raw slopes were winsorized at the 1st and 99th percentiles of their empirical distribution.

Baseline BMI, systolic blood pressure, and diastolic blood pressure were ascertained from the 90 days preceding the index date. Comorbid conditions and overall comorbidity burden, summarized with the age-adjusted Charlson Comorbidity Index, were identified from ICD-10 diagnosis codes and structured medication orders recorded before the index date (**eTable 1**).

**Regression-Adjusted Absolute Differences by Race and Ethnicity, Social Vulnerability Index, and Insurance Type**

To evaluate whether subgroup differences in 12-month prescribing persistence, attainment of the maintenance dose by 12 months, and ≥5% TBWL reflected confounding by geography or calendar time, we fit multivariable logistic regression models for each outcome, separately for semaglutide and tirzepatide. Race/ethnicity, SVI quartile, and insurance type were included together in each model (mutually adjusted), along with state of residence and calendar month of treatment initiation. Reference categories were non-Hispanic White, SVI quartile 1, and commercial insurance. Adjusted absolute differences for each subgroup level, relative to its reference category, were estimated by marginal standardization: each patient's exposure level was set to the level of interest while all other covariates were held at their observed values, the model-predicted probability was averaged across all patients, and the difference from the analogous average under the reference level was taken as the adjusted absolute difference. 95% confidence intervals were obtained by nonparametric bootstrap with patient-level resampling and model refitting (200 replicates).

**Supplement Figure 1. Flow Chart of Participants.**

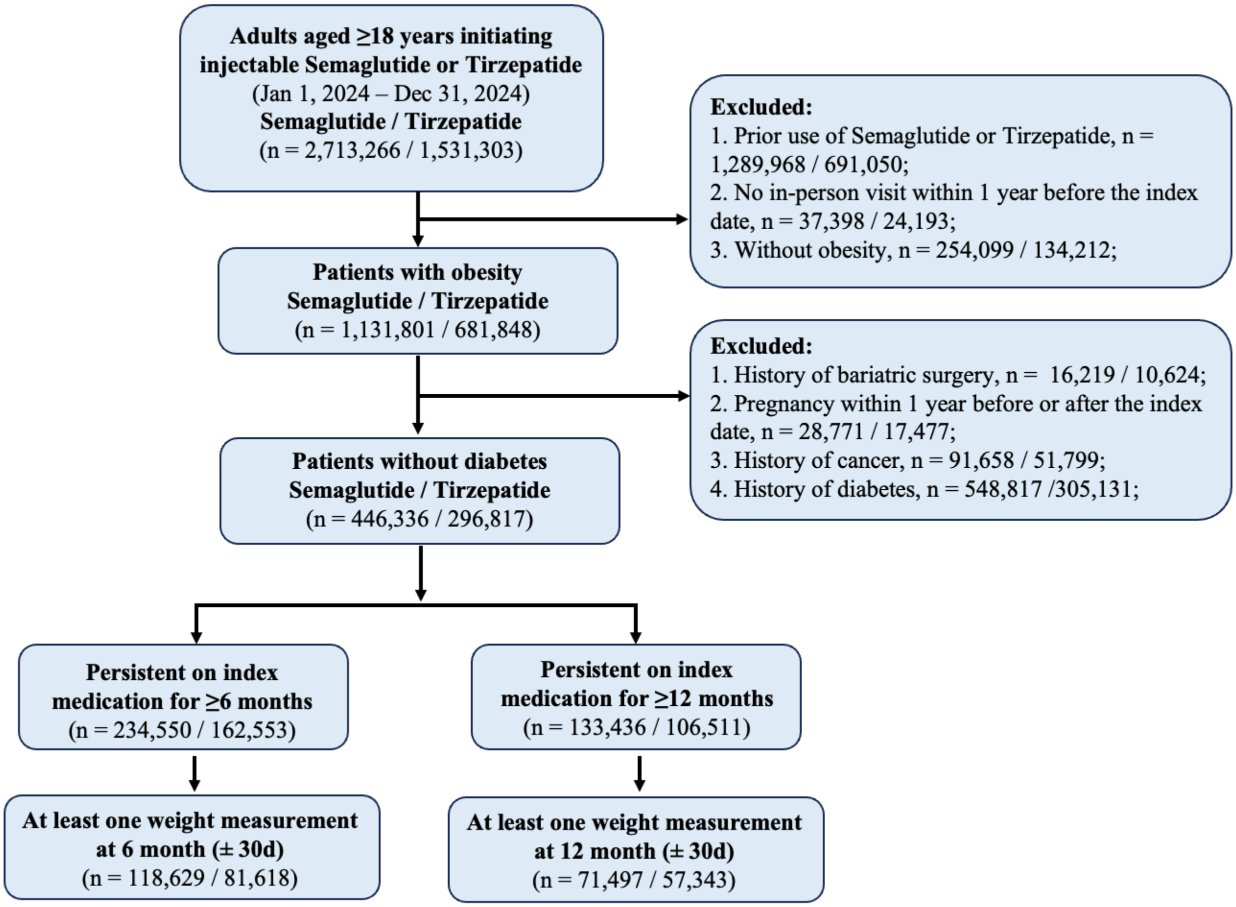

**Supplement Figure 2. Post-Initiation Care Pattern from Treatment Initiation To 12 Months Among Patients Initiating Semaglutide or Tirzepatide.**

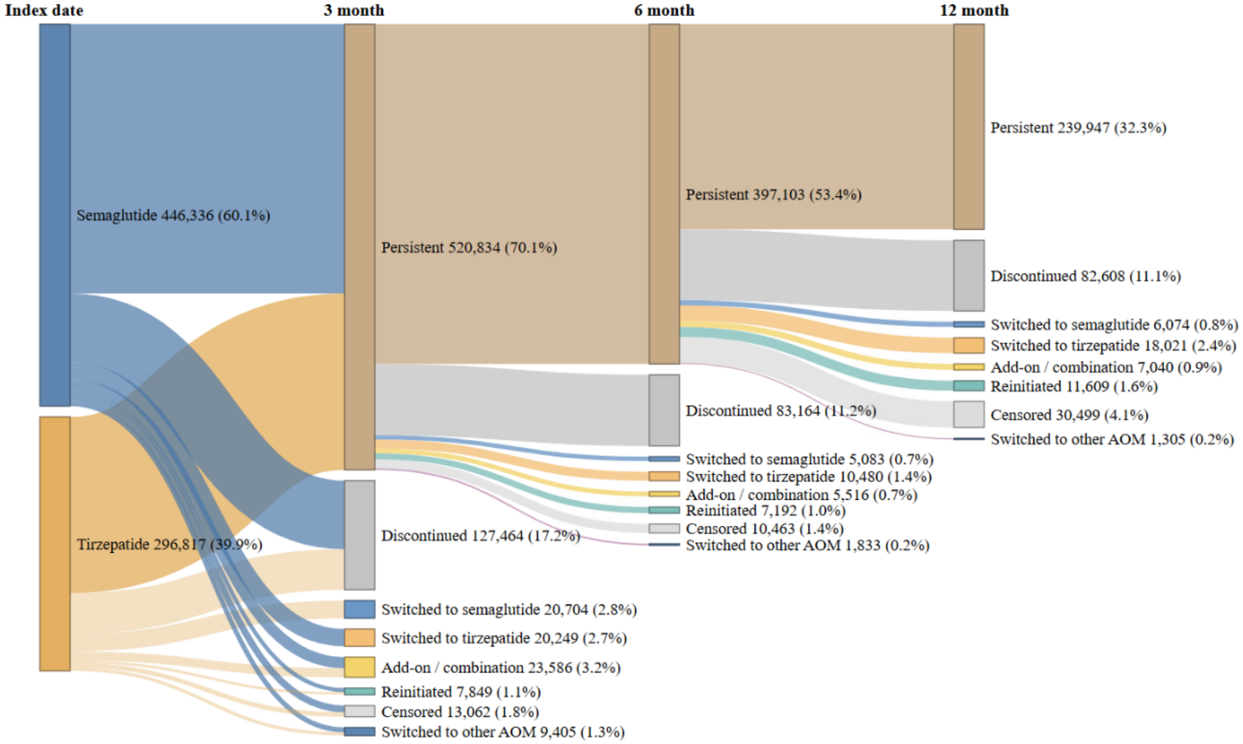


Shown is a Sankey diagram of the combined post-initiation care pattern among all semaglutide and tirzepatide initiators, using the same hierarchical categorization and 60-day gap, with semaglutide and tirzepatide initiators combined into a single cohort at each landmark.

**Supplement Figure 3. Sensitivity Analysis of Post-Initiation Care Pattern Using A 30-Day Gap Between Prescription Orders.**

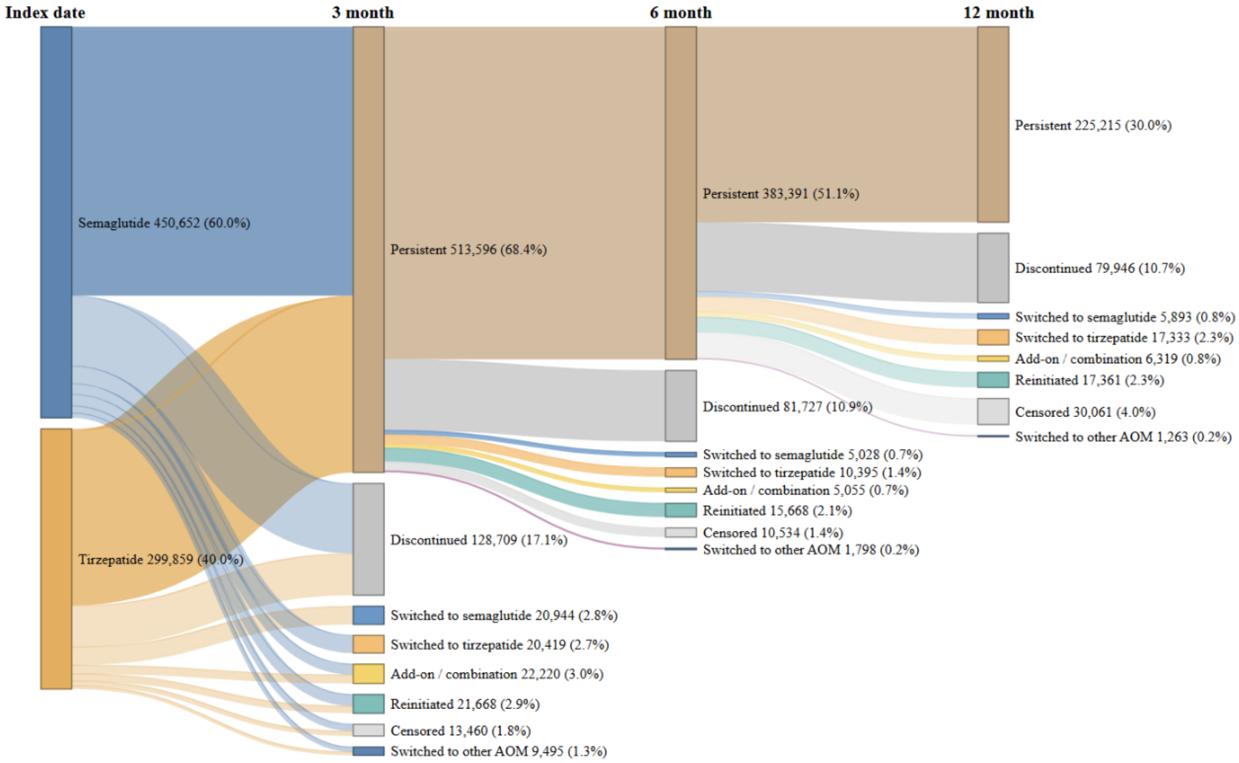


Shown is the post-initiation care pattern with prescribing persistence defined using a maximum allowable gap of 30 days (rather than 60 days) between consecutive prescription orders for the index medication.

**Supplement Figure 4. Sensitivity Analysis of Post-Initiation Care Pattern Using A 90-Day Gap Between Prescription Orders.**

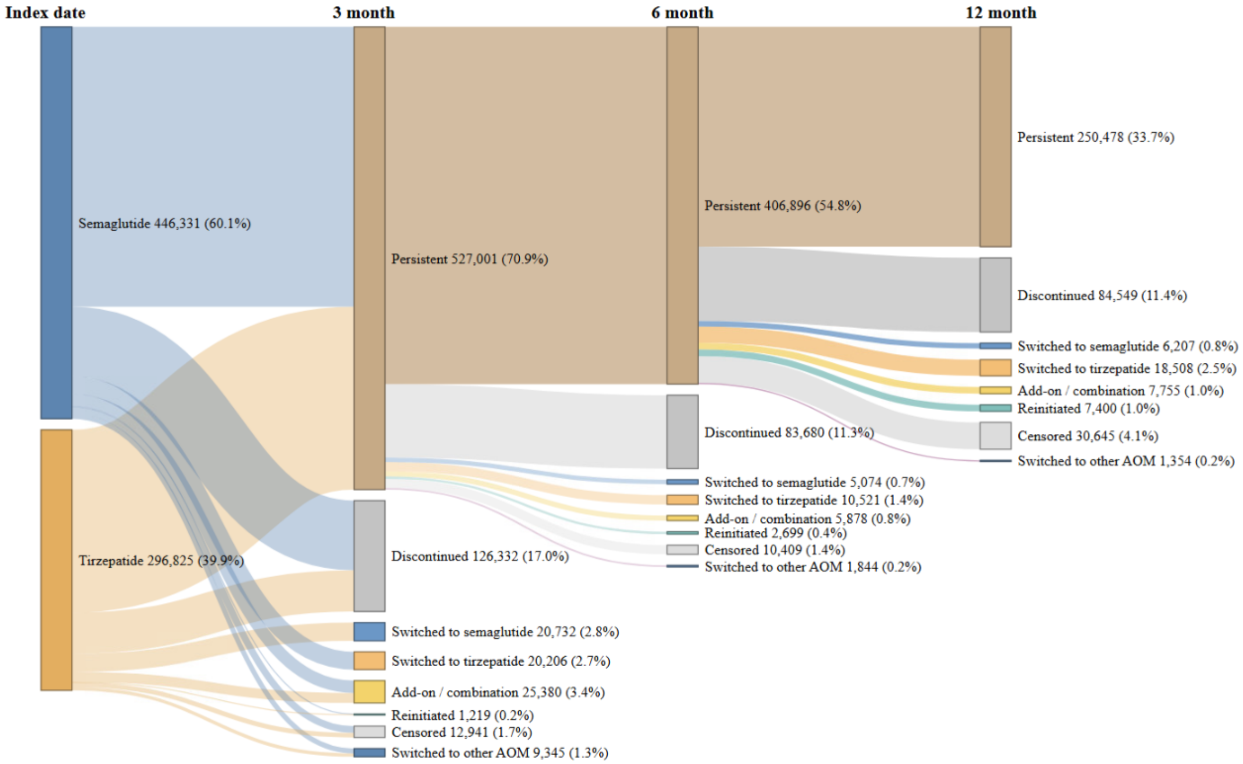


Shown is the post-initiation care pattern with prescribing persistence defined using a maximum allowable gap of 90 days (rather than 60 days) between consecutive prescription orders for the index medication.

**Supplement Figure 5. Sensitivity Analysis of Post-Initiation Care Pattern Among Patients Initiating WEGOVY or ZEPBOUND.**

**(A) WEGOVY**

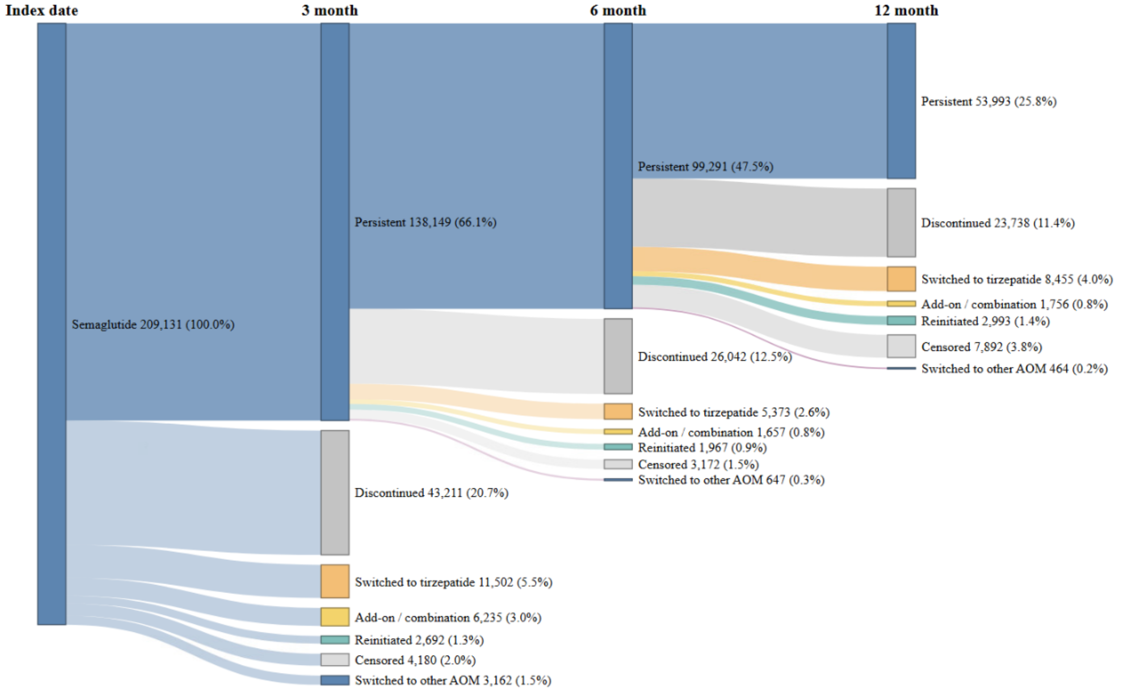


**(B) ZEPBOUND**

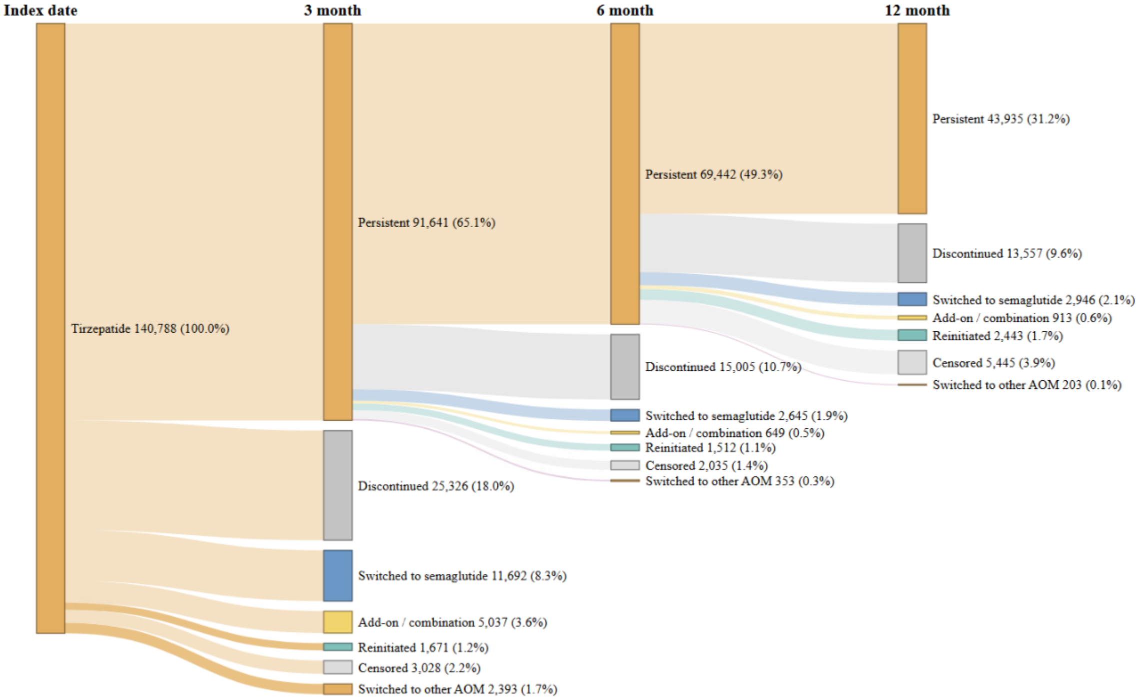


Shown is the post-initiation care pattern restricted to initiators of (A) WEGOVY or (B) ZEPBOUND, respectively.

**Supplement Figure 6. Stepwise Attrition Along the Post-Initiation Care Continuum Among Semaglutide Initiators, Overall and by Race/Ethnicity, Social Vulnerability Index Quartile, and Insurance Type.**

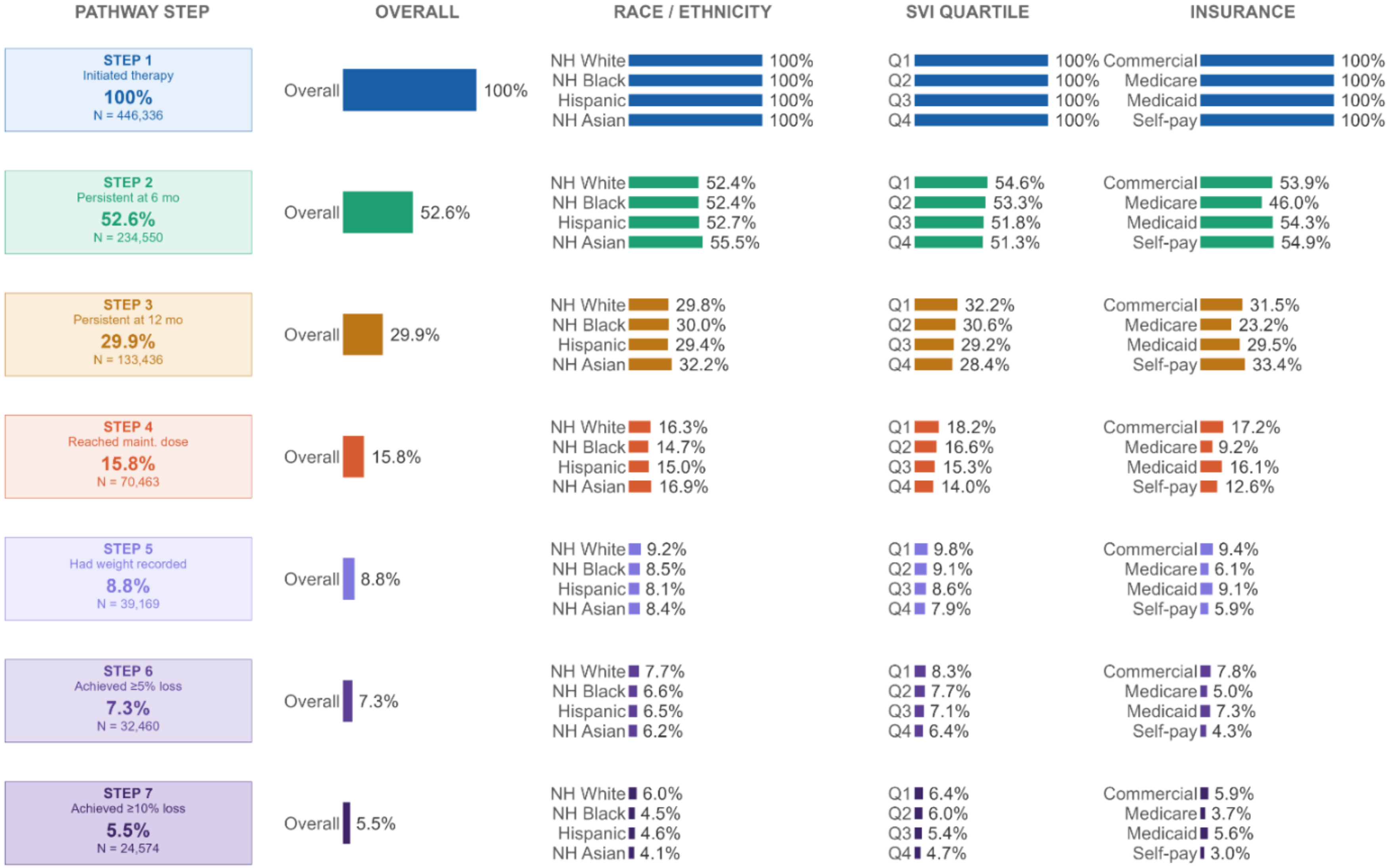


Shown is the proportion of initiators remaining at each step of the post-initiation care cascade overall and stratified by race/ethnicity, Social Vulnerability Index (SVI) quartile, and insurance type, among initiators of semaglutide.

**Supplement Figure 7. Stepwise Attrition Along the Post-Initiation Care Continuum Among Tirzepatide Initiators, Overall and by Race/Ethnicity, Social Vulnerability Index Quartile, and Insurance Type.**

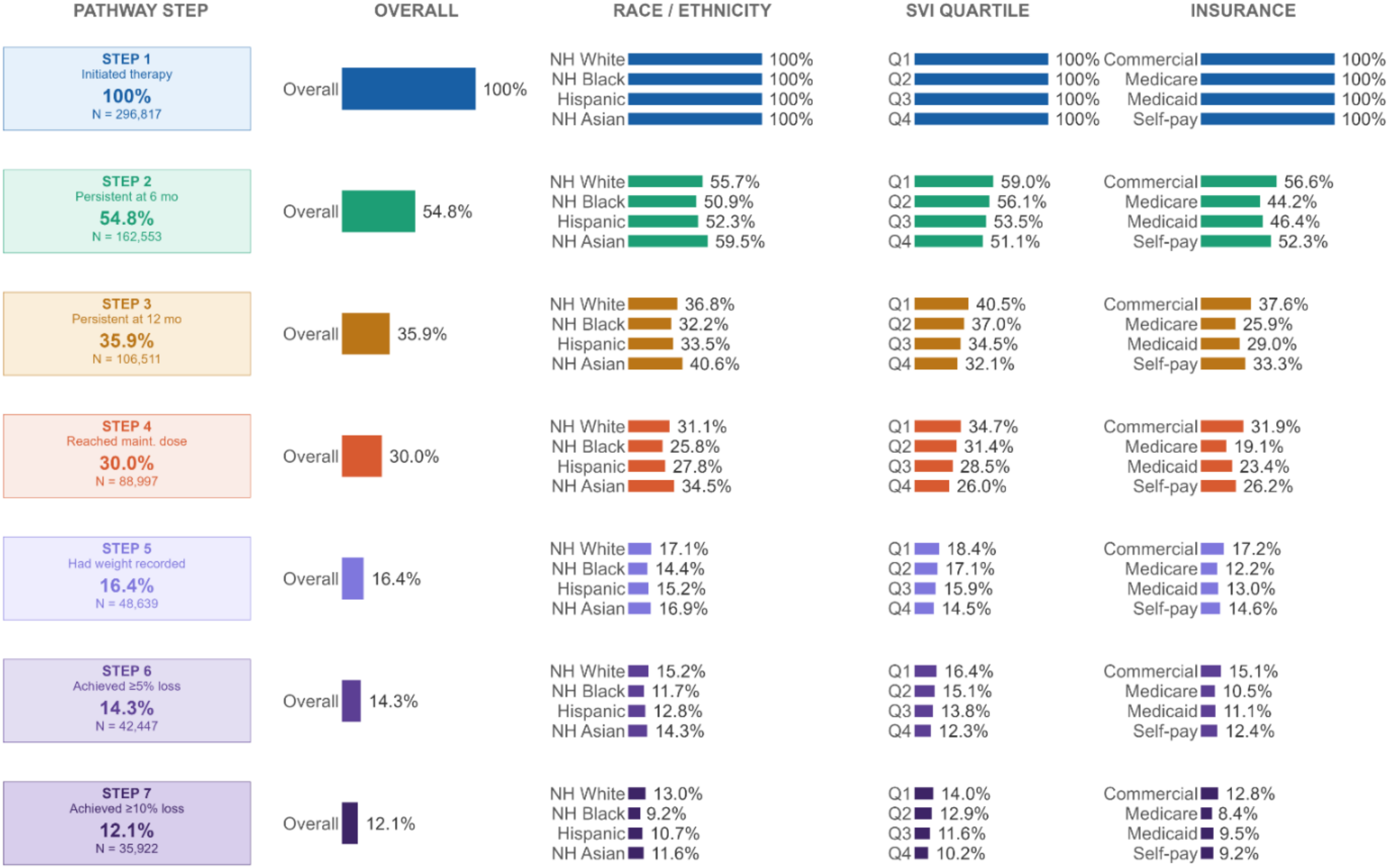


Shown is the proportion of initiators remaining at each step of the post-initiation care cascade overall and stratified by race/ethnicity, Social Vulnerability Index (SVI) quartile, and insurance type, among initiators of tirzepatide.

**Supplement Figure 8. Stepwise Attrition Along the Post-Initiation Care Continuum Among WEGOVY Initiators, Overall and by Race/Ethnicity, Social Vulnerability Index Quartile, and Insurance Type.**

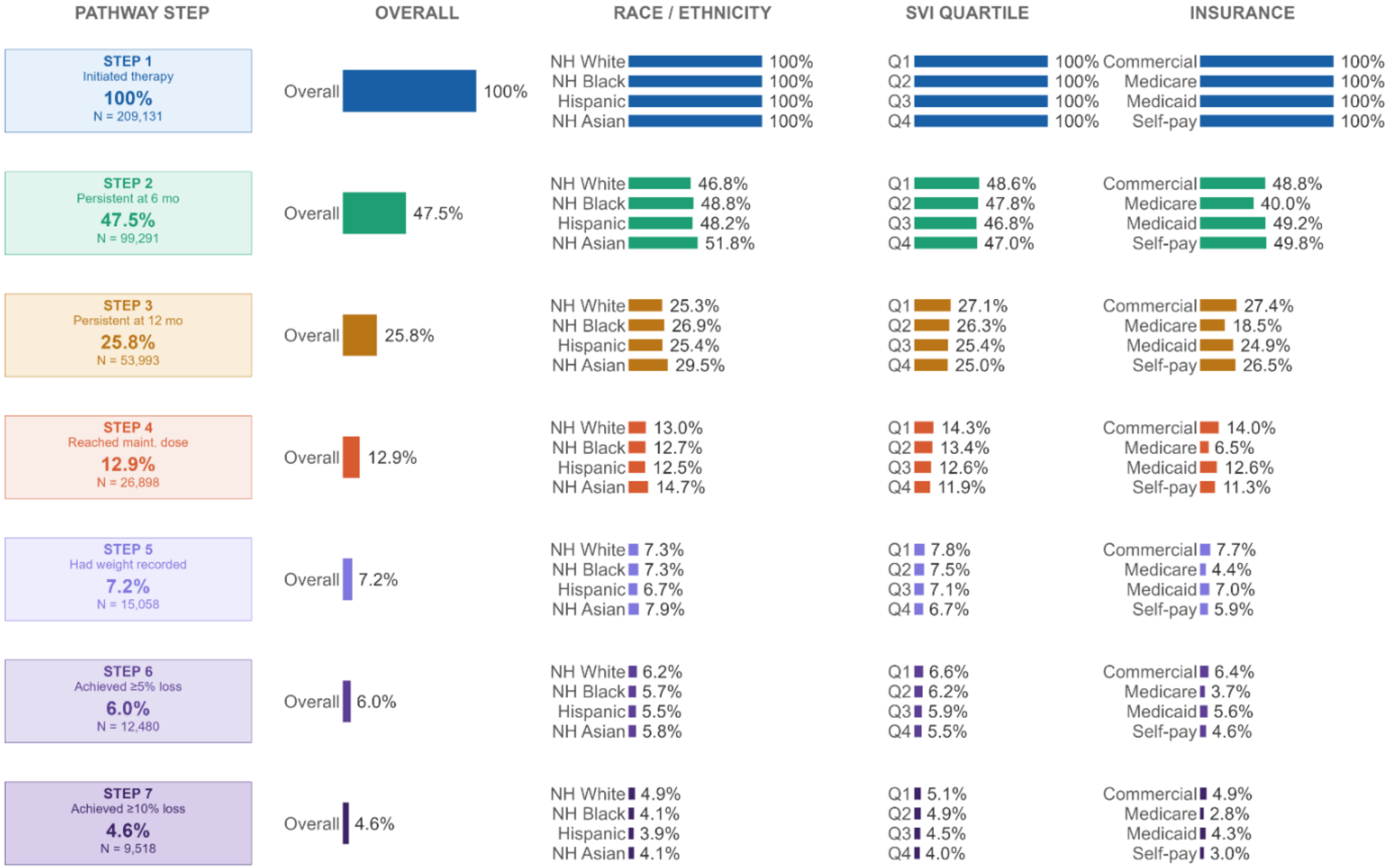


Shown is the proportion of initiators remaining at each step of the post-initiation care cascade overall and stratified by race/ethnicity, Social Vulnerability Index (SVI) quartile, and insurance type, among initiators of WEGOVY.

**Supplement Figure 9. Stepwise Attrition Along the Post-Initiation Care Continuum Among ZEPBOUND Initiators, Overall and by Race/Ethnicity, Social Vulnerability Index Quartile, and Insurance Type.**

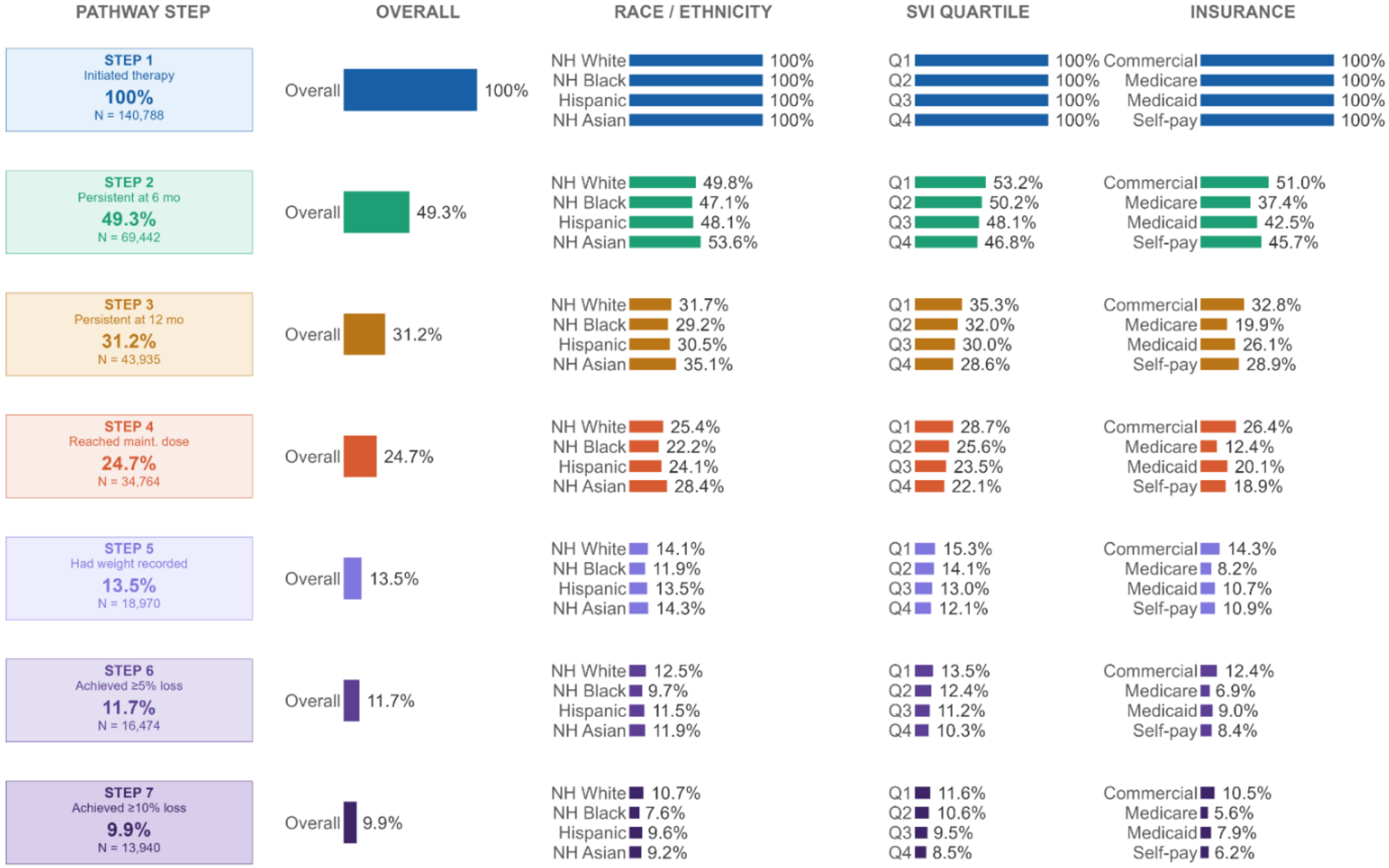


Shown is the proportion of initiators remaining at each step of the post-initiation care cascade overall and stratified by race/ethnicity, Social Vulnerability Index (SVI) quartile, and insurance type, among initiators of ZEPBOUND.

**Supplement Figure 10. FDA-Labeled Maintenance-Dose Attainment and Monthly Dose Distribution Over 12 Months Among Initiators Prescribing Persistent Through 12 Months.**

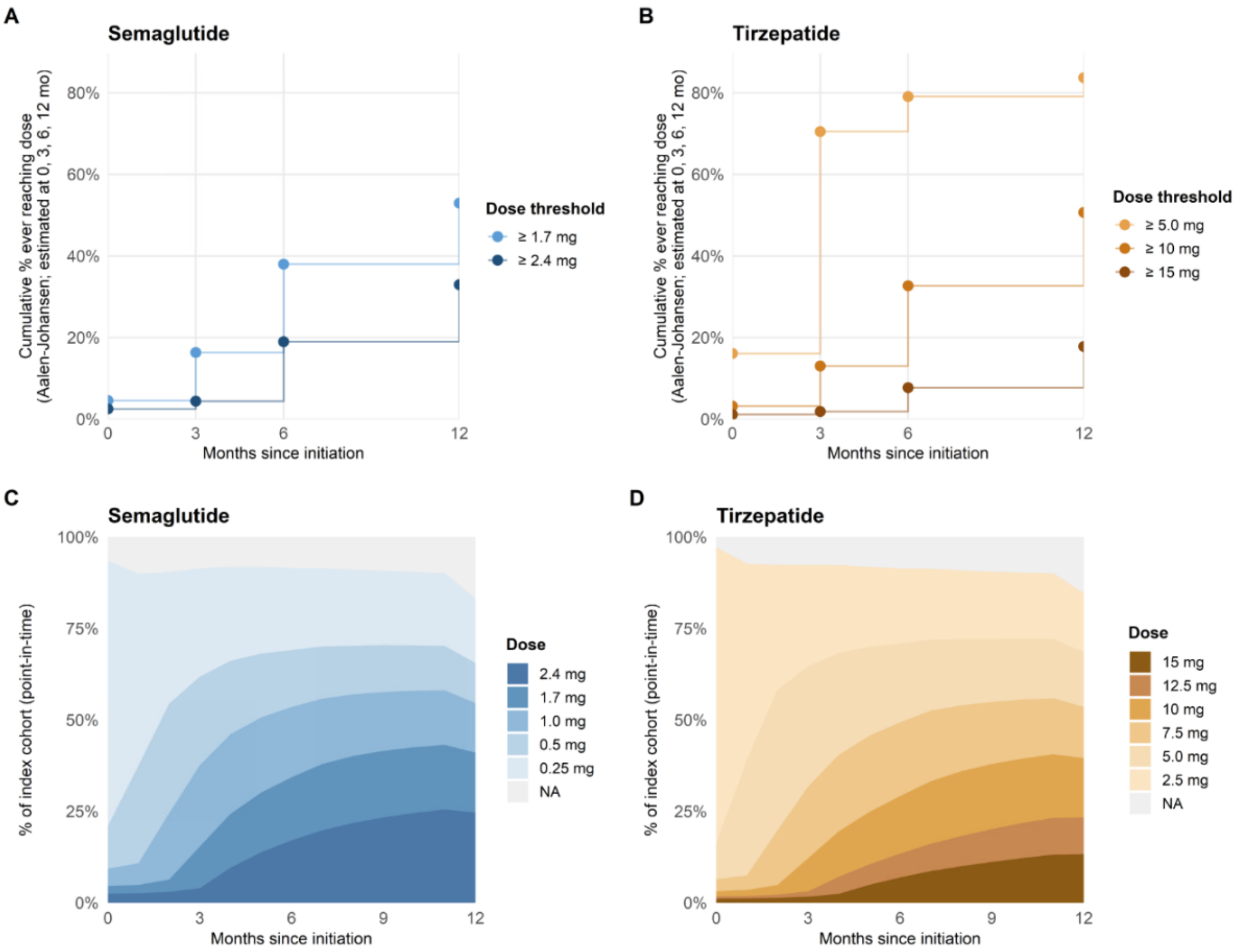


(A, B) Cumulative incidence of ever reaching each weekly dose threshold for semaglutide (≥1.7 mg, the initial FDA-labeled maintenance dose, and ≥2.4 mg) and tirzepatide (≥5 mg, the initial FDA-labeled maintenance dose, and ≥10 mg and ≥15 mg).
(C, D) Point-in-time distribution of the prescribed weekly dose among initiators prescribing persistent through 12 months at monthly intervals after initiation. NA indicates no active order (including after discontinuation) or missing dose.
FDA indicates Food and Drug Administration.

**Supplement Figure 11. Cumulative Incidence of Reaching the Initial FDA-Labeled Maintenance Dose (Semaglutide: 1.7 mg; Tirzepatide: 5 mg) by Subgroups.**

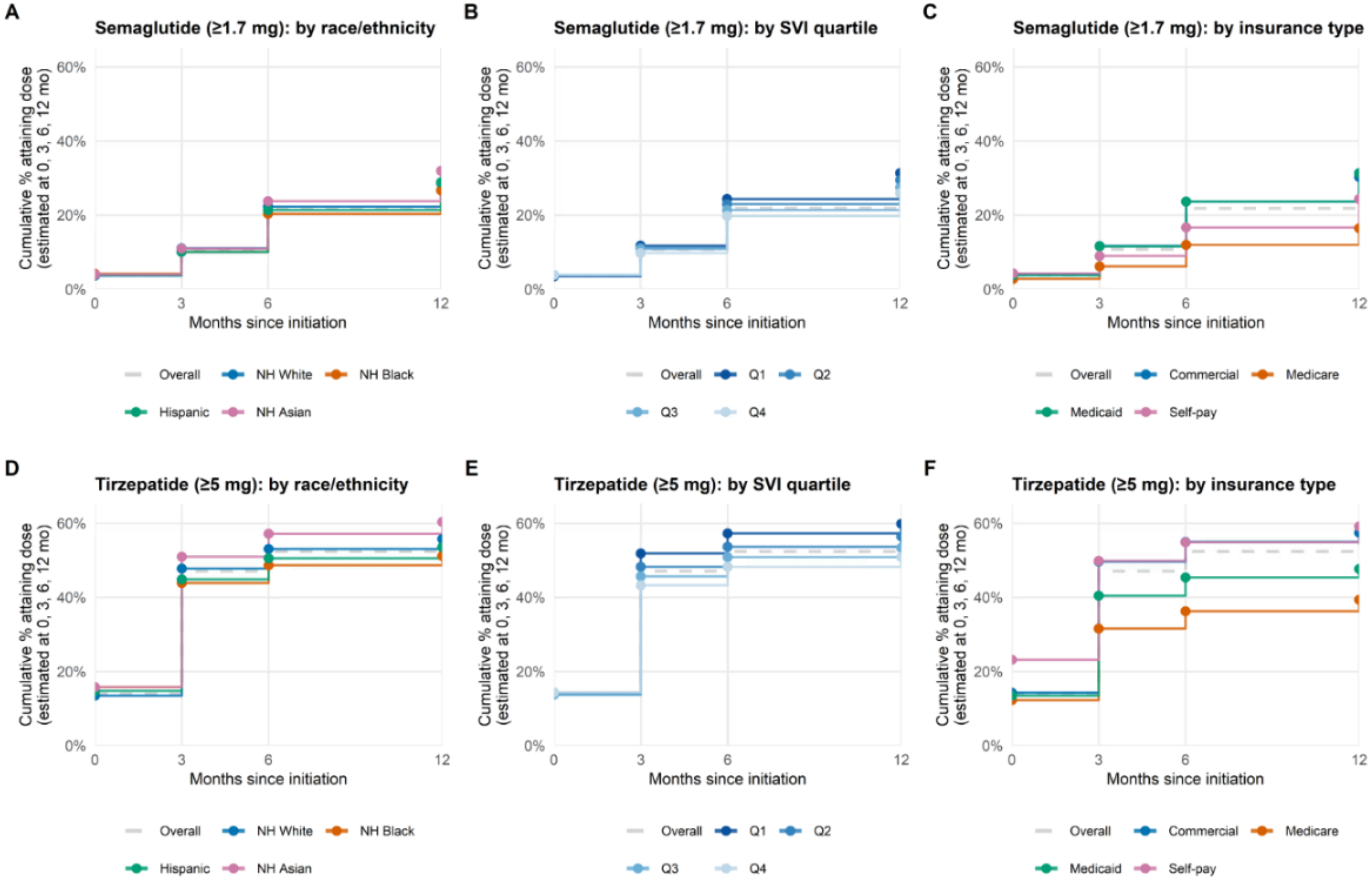


Shown are cumulative incidence curves, estimated with the Aalen-Johansen method, for attainment of the initial FDA-labeled maintenance dose, stratified by race/ethnicity, Social Vulnerability Index (SVI) quartile, and insurance type, among semaglutide and tirzepatide initiators.

**Supplement Figure 12. Cumulative Incidence of Reaching the Higher FDA-Labeled Maintenance Dose (Semaglutide: 2.4 mg; Tirzepatide: 15 mg) by Subgroups.**

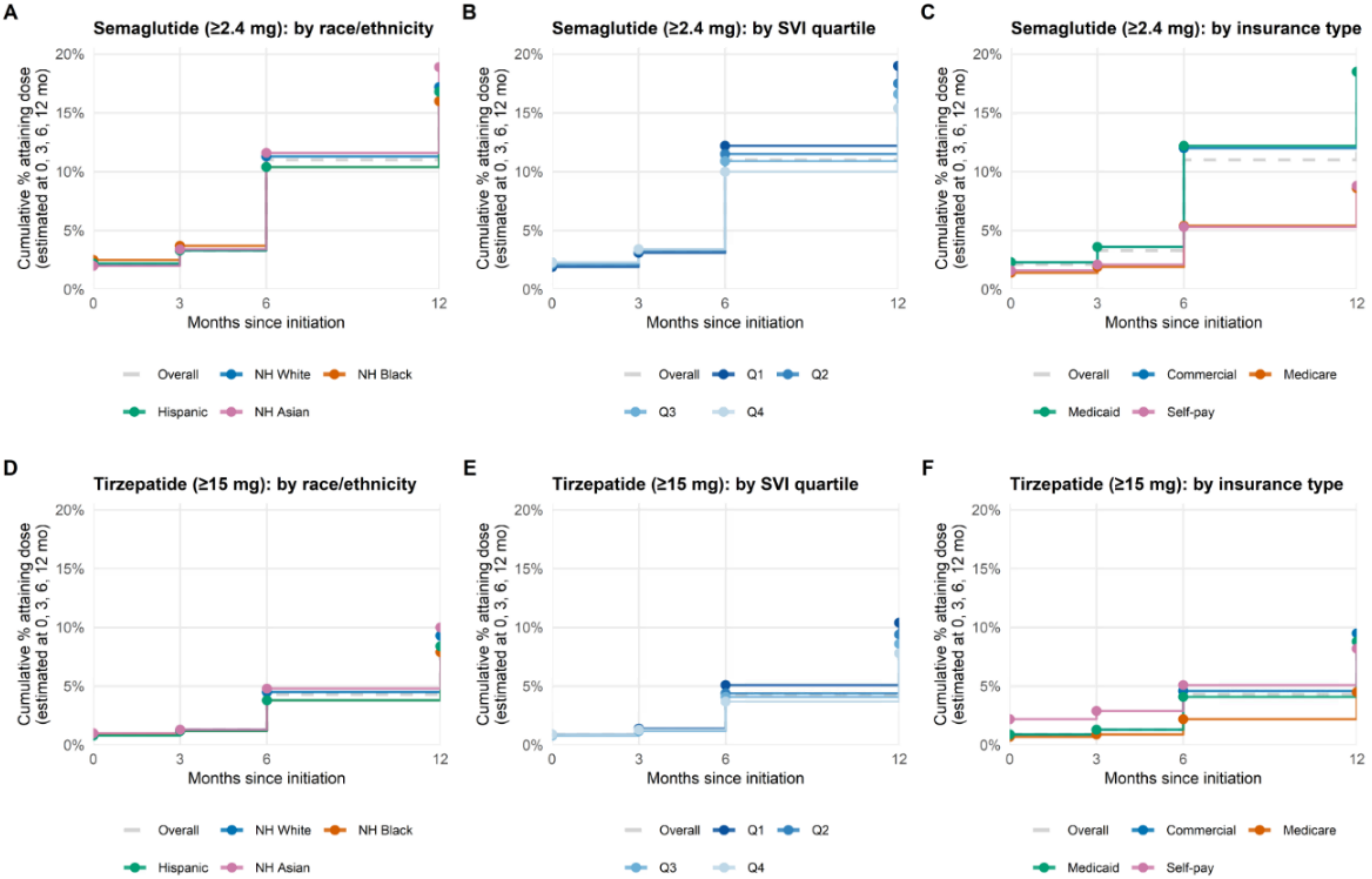


Shown are cumulative incidence curves, estimated with the Aalen-Johansen method, for attainment of the higher FDA-labeled maintenance dose, stratified by race/ethnicity, Social Vulnerability Index (SVI) quartile, and insurance type, among semaglutide and tirzepatide initiators.

**Supplement Figure 13. Weight Change Among Adults Initiating Semaglutide or Tirzepatide for Obesity At 6 Months.**

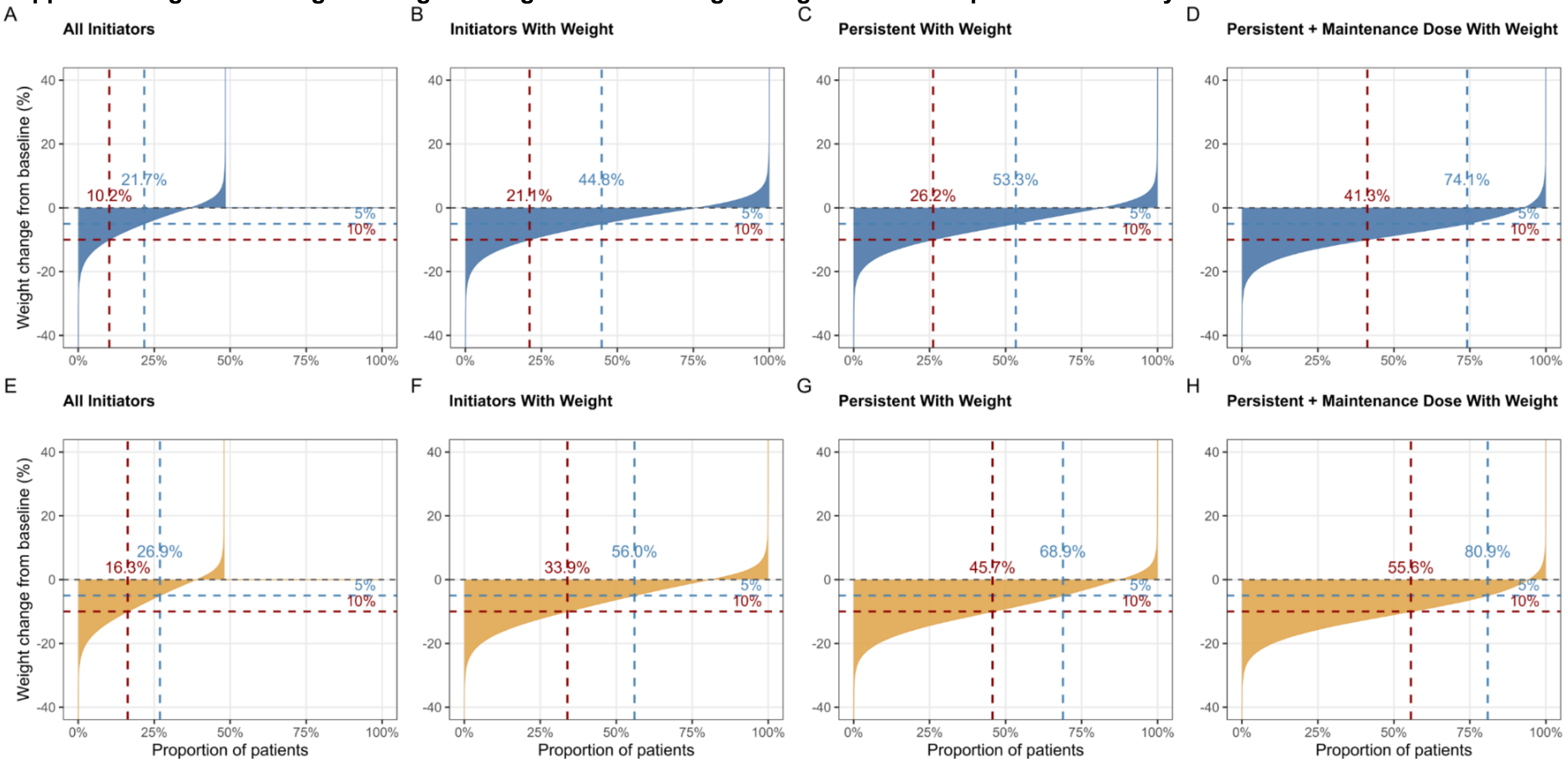


Shown is the distribution of percent total body weight change from baseline at 6 months among (A, E) all initiators, (B, F) initiators with a recorded weight, (C, G) initiators who were prescribing persistent with a recorded weight, and (D, H) initiators who were prescribing persistent at an FDA-labeled maintenance dose with a recorded weight, for (A-D) semaglutide and (E-H) tirzepatide. Each curve represents the distribution of percent weight change from baseline, with patients ordered from greatest to least weight loss; percentages shown indicate the proportion of patients achieving at least 5% (blue) or 10% (red) weight loss.

**Supplement Table 1. Definitions of Study Variables and Corresponding Codes.**

| Conditions | ICD-10 Codes | CPT Codes |
|---|---|---|
| Diabetes | E08, E09, E10, E11, E13 | / |
| Hypertension | I10, I11, I12, I13, I15 | / |
| Dyslipidemia | E78.0- E78.6 | / |
| Obstructive sleep apnea | G47.33 | |
| Pregnancy | O00-O9A | |
| Cancer | C00-C96 | |
| **Bariatric Surgery** | | |
| Biliopancreatic diversion | / | 43845 |
| Gastric banding | / | 43770 |
| Roux-en-Y | / | 43633, 43644, 43645, 43846, 43847 |
| Sleeve gastrectomy | / | 43775, 43842, 43843 |
| Other gastrectomy | / | 43659 |
| **Anti-obesity Medications (Index/Comparator)** | Semaglutide, tirzepatide (injectable formulations) | |
| **Other Anti-obesity Medications** | Liraglutide; orlistat; phentermine HCl; phentermine resin; phentermine/topiramate; naltrexone HCl/bupropion HCl; sibutramine HCl m-hydrate; phenylpropanolamine HCl; mazindol; lorcaserin HCl; phendimetrazine tartrate | |
| **Anti-Diabetic Medications** | | |
| Biguanide | Metformin | |
| Insulin | Insulin (all formulations) | |
| DPP-4 Inhibitor | Generic name containing "gliptin" | |
| GLP-1 Receptor Agonist (Other) | Albiglutide; dulaglutide; exenatide; lixisenatide; oral semaglutide | |
| SGLT2 Inhibitor | Generic name containing "gliflozin" | |
| Sulfonylurea | Chlorpropamide; glimepiride; glipizide; gliquidone; glyburide; tolazamide; tolbutamide | |
| Alpha-Glucosidase Inhibitor | Acarbose | |
| **Items for ACCI** | | |
| Myocardial infarction | I21, I22, I252 | |
| Congestive heart failure | I43, I50, I099, I110, I130, I132, I255, I420, I425, I426, I427, I428, I429, P290 | |
| Peripheral vascular disease | I70, I71, I731, I738, I739, I771, I790, I792, K551, K558, K559, Z958, Z959 | |
| Cerebrovascular disease | G45, G46, H340, I60, I61, I62, I63, I64, I65, I66, I67, I68, I69 | |
| Dementia | F00, F01, F02, F03, F051, G30, G311 | |
| Chronic pulmonary disease | I278, I279, J40, J41, J42, J43, J44, J45, J46, J47, J60, J61, J62, J63, J64, J65, J66, J67, J684, J701, J703 | |
| Rheumatic disease | M05, M06, M32, M33, M34, M315, M351, M353, M360 | |
| Peptic ulcer disease | K25, K26, K27, K28 | |
| Mild liver disease | B18, K700, K701, K702, K703, K709, K713, K714, K715, K717, K73, K74, K760, K762, K763, K764, K768, K769, Z944 | |
| Diabetes without complications | E100, E101, E106, E108, E109, E110, E111, E116, E118, E119, E120, E121, E126, E128, E129, E130, E131, E136, E138, E139, E140, E141, E146, E148, E149 | |
| Diabetes with complications | E102, E103, E104, E105, E107, E112, E113, E114, E115, E117, E122, E123, E124, E125, E127, E132, E133, E134, E135, E137, E142, E143, E144, E145, E147 | |

| | |
|---|---|
| Hemiplegia or paraplegia | G041, G114, G801, G802, G81, G82, G830, G831, G832, G833, G834, G839 |
| Moderate or severe renal disease | I120, I131, N032, N033, N034, N035, N036, N037, N052, N053, N054, N055, N056, N057, N18, N19, N250, Z490, Z491, Z492, Z940, Z992 |
| Any malignancy (excluding metastatic) | C0, C1, C2, C30, C31, C32, C33, C34, C37, C38, C39, C40, C41, C43, C45, C46, C47, C48, C49, C50, C51, C52, C53, C54, C55, C56, C57, C58, C6, C70, C71, C72, C73, C74, C75, C76, C81, C82, C83, C84, C85, C88, C90, C91, C92, C93, C94, C95, C96, C97 |
| Severe liver disease | I850, I859, I864, I982, K704, K711, K721, K729, K765, K766, K767 |
| Metastatic solid tumor | C77, C78, C79, C80 |
| AIDS/HIV | B20, B21, B22, B24 |

Comorbid conditions were ascertained using diagnosis codes recorded before the index date;
ACCI denotes age-adjusted Charlson Comorbidity Index; AIDS denotes acquired immunodeficiency syndrome; CPT denotes Current Procedural Terminology; HIV denotes human immunodeficiency virus; ICD-10 denotes International Classification of Diseases, Tenth Revision.

**Supplement Table 2. Post-Initiation Care Over 12 Months With 95% Confidence Intervals.**

| | 3-Month | 6-Month | 12-Month |
|---|---|---|---|
| **Semaglutide** | | | |
| Persistent | 70.5 (70.4-70.7) | 52.6 (52.4-52.7) | 29.9 (29.8-30.0) |
| Discontinued | 17.9 (17.8-18.1) | 12.0 (11.9-12.1) | 11.9 (11.8-12.0) |
| Switched to tirzepatide | 4.5 (4.5-4.6) | 2.3 (2.3-2.4) | 4.0 (4.0-4.1) |
| Add-on / combination | 2.9 (2.9-3.0) | 0.9 (0.9-0.9) | 1.0 (1.0-1.0) |
| Reinitiated | 1.1 (1.0-1.1) | 0.9 (0.9-1.0) | 1.4 (1.4-1.5) |
| Censored | 1.7 (1.7-1.8) | 1.5 (1.4-1.5) | 4.1 (4.1-4.2) |
| Switched to other AOM | 1.3 (1.2-1.3) | 0.3 (0.3-0.3) | 0.2 (0.2-0.2) |
| **Tirzepatide** | | | |
| Persistent | 69.4 (69.3-69.6) | 54.8 (54.6-54.9) | 35.9 (35.7-36.1) |
| Discontinued | 16.0 (15.8-16.1) | 9.9 (9.8-10.0) | 10.0 (9.9-10.1) |
| Switched to semaglutide | 7.0 (6.9-7.1) | 1.7 (1.7-1.8) | 2.0 (2.0-2.1) |
| Add-on / combination | 3.5 (3.5-3.6) | 0.5 (0.5-0.6) | 0.9 (0.8-0.9) |
| Reinitiated | 1.0 (1.0-1.1) | 1.0 (1.0-1.0) | 1.8 (1.7-1.8) |
| Censored | 1.8 (1.7-1.8) | 1.3 (1.3-1.3) | 4.1 (4.0-4.2) |
| Switched to other AOM | 1.3 (1.2-1.3) | 0.2 (0.2-0.2) | 0.1 (0.1-0.1) |
| **Overall** | | | |
| Persistent | 70.1 (70.0-70.2) | 53.4 (53.3-53.5) | 32.3 (32.2-32.4) |
| Discontinued | 17.2 (17.1-17.2) | 11.2 (11.1-11.3) | 11.1 (11.0-11.2) |
| Switched to semaglutide | 2.8 (2.7-2.8) | 0.7 (0.7-0.7) | 0.8 (0.8-0.8) |
| Switched to tirzepatide | 2.7 (2.7-2.8) | 1.4 (1.4-1.4) | 2.4 (2.4-2.5) |
| Add-on / combination | 3.2 (3.1-3.2) | 0.7 (0.7-0.8) | 0.9 (0.9-1.0) |
| Reinitiated | 1.1 (1.0-1.1) | 1.0 (0.9-1.0) | 1.6 (1.5-1.6) |
| Censored | 1.8 (1.7-1.8) | 1.4 (1.4-1.4) | 4.1 (4.1-4.1) |
| Switched to other AOM | 1.3 (1.2-1.3) | 0.2 (0.2-0.3) | 0.2 (0.2-0.2) |

Values are percentages (95% confidence interval) of all initiators of each medication.

**Supplement Table 3. Sensitivity Analyses of Post-Initiation Care Over 12 Months With 95% Confidence Intervals.**

| | 3-Month | 6-Month | 12-Month |
|---|---|---|---|
| **Using A 30-Day Gap** | | | |
| Persistent | 68.4 (68.3-68.5) | 51.1 (51.0-51.2) | 30.0 (29.9-30.1) |
| Discontinued | 17.1 (17.1-17.2) | 10.9 (10.8-11.0) | 10.7 (10.6-10.7) |
| Switched to semaglutide | 2.8 (2.8-2.8) | 0.7 (0.7-0.7) | 0.8 (0.8-0.8) |
| Switched to tirzepatide | 2.7 (2.7-2.8) | 1.4 (1.4-1.4) | 2.3 (2.3-2.3) |
| Add-on / combination | 3.0 (2.9-3.0) | 0.7 (0.7-0.7) | 0.8 (0.8-0.9) |
| Reinitiated | 2.9 (2.8-2.9) | 2.1 (2.1-2.1) | 2.3 (2.3-2.3) |
| Censored | 1.8 (1.8-1.8) | 1.4 (1.4-1.4) | 4.0 (4.0-4.1) |
| Switched to other AOM | 1.3 (1.2-1.3) | 0.2 (0.2-0.3) | 0.2 (0.2-0.2) |
| **Using A 90-Day Gap** | | | |
| Persistent | 70.9 (70.8-71.0) | 54.8 (54.6-54.9) | 33.7 (33.6-33.8) |
| Discontinued | 17.0 (16.9-17.1) | 11.3 (11.2-11.3) | 11.4 (11.3-11.4) |
| Switched to semaglutide | 2.8 (2.8-2.8) | 0.7 (0.7-0.7) | 0.8 (0.8-0.9) |
| Switched to tirzepatide | 2.7 (2.7-2.8) | 1.4 (1.4-1.4) | 2.5 (2.5-2.5) |
| Add-on / combination | 3.4 (3.4-3.5) | 0.8 (0.8-0.8) | 1.0 (1.0-1.1) |
| Reinitiated | 0.2 (0.2-0.2) | 0.4 (0.3-0.4) | 1.0 (1.0-1.0) |
| Censored | 1.7 (1.7-1.8) | 1.4 (1.4-1.4) | 4.1 (4.1-4.2) |
| Switched to other AOM | 1.3 (1.2-1.3) | 0.2 (0.2-0.3) | 0.2 (0.2-0.2) |
| **WEGOVY** | | | |
| Persistent | 66.1 (65.9-66.3) | 47.5 (47.3-47.7) | 25.8 (25.6-26.0) |
| Discontinued | 20.7 (20.5-20.8) | 12.5 (12.3-12.6) | 11.4 (11.2-11.5) |
| Switched to tirzepatide | 5.5 (5.4-5.6) | 2.6 (2.5-2.6) | 4.0 (4.0-4.1) |
| Add-on / combination | 3.0 (2.9-3.1) | 0.8 (0.8-0.8) | 0.8 (0.8-0.9) |
| Reinitiated | 1.3 (1.2-1.3) | 0.9 (0.9-1.0) | 1.4 (1.4-1.5) |
| Censored | 2.0 (1.9-2.1) | 1.5 (1.4-1.5) | 3.8 (3.7-3.9) |
| Switched to other AOM | 1.5 (1.5-1.6) | 0.3 (0.3-0.3) | 0.2 (0.2-0.2) |
| **ZEPBOUND** | | | |
| Persistent | 65.1 (64.8-65.3) | 49.3 (49.1-49.6) | 31.2 (31.0-31.4) |
| Discontinued | 18 (17.8-18.2) | 10.7 (10.5-10.8) | 9.6 (9.5-9.8) |
| Switched to semaglutide | 8.3 (8.2-8.4) | 1.9 (1.8-2.0) | 2.1 (2.0-2.2) |
| Add-on / combination | 3.6 (3.5-3.7) | 0.5 (0.4-0.5) | 0.6 (0.6-0.7) |
| Reinitiated | 1.2 (1.1-1.2) | 1.1 (1.0-1.1) | 1.7 (1.7-1.8) |
| Censored | 2.2 (2.1-2.2) | 1.4 (1.4-1.5) | 3.9 (3.8-4.0) |
| Switched to other AOM | 1.7 (1.6-1.8) | 0.3 (0.2-0.3) | 0.1 (0.1-0.2) |
| **WEGOVY+ ZEPBOUND** | | | |
| Persistent | 65.7 (65.5-65.8) | 48.2 (48.1-48.4) | 28 (27.8-28.1) |
| Discontinued | 19.6 (19.5-19.7) | 11.7 (11.6-11.8) | 10.7 (10.6-10.8) |
| Switched to semaglutide | 3.3 (3.3-3.4) | 0.8 (0.7-0.8) | 0.8 (0.8-0.9) |
| Switched to tirzepatide | 3.3 (3.2-3.3) | 1.5 (1.5-1.6) | 2.4 (2.4-2.5) |
| Add-on / combination | 3.2 (3.2-3.3) | 0.7 (0.6-0.7) | 0.8 (0.7-0.8) |
| Reinitiated | 1.2 (1.2-1.3) | 1.0 (1.0-1.0) | 1.6 (1.5-1.6) |
| Censored | 2.1 (2.0-2.1) | 1.5 (1.4-1.5) | 3.8 (3.7-3.9) |
| Switched to other AOM | 1.6 (1.5-1.6) | 0.3 (0.2-0.3) | 0.2 (0.2-0.2) |

Values are percentages (95% confidence interval) of all initiators, using alternative persistence-gap definitions (30-day and 90-day allowable gaps between consecutive prescription orders) and restricted to initiators of WEGOVY, ZEPBOUND, and both combined.

**Supplement Table 4. Weight-Loss Outcomes of Semaglutide Initiators by Care Pattern.**

| | N | Denominator | PCT (%) | 95% CI Low | 95% CI High |
|---|---|---|---|---|---|
| **3-Month Weight Outcome (Non-persistent by 3m, n = 131,583)** | | | | | |
| ≥5% | 14,036 | 131,583 | 10.7 | 10.5 | 10.8 |
| ≥10% | 3,247 | 131,583 | 2.5 | 2.4 | 2.6 |
| 5-10% | 10,789 | 131,583 | 8.2 | 8.1 | 8.3 |
| <5% | 47,190 | 131,583 | 35.9 | 35.6 | 36.1 |
| No wt | 70,357 | 131,583 | 53.5 | 53.2 | 53.7 |
| **6-Month Weight Outcome (Non-persistent by 6m, n = 80,203)** | | | | | |
| ≥5% | 12,395 | 80,203 | 15.5 | 15.2 | 15.7 |
| ≥10% | 4,926 | 80,203 | 6.1 | 6.0 | 6.3 |
| 5-10% | 7,469 | 80,203 | 9.3 | 9.1 | 9.5 |
| <5% | 25,640 | 80,203 | 32.0 | 31.6 | 32.3 |
| No wt | 42,168 | 80,203 | 52.6 | 52.2 | 52.9 |
| **12-Month Weight Outcome (Non-persistent by 12m, n = 101,114)** | | | | | |
| ≥5% | 23,151 | 101,114 | 22.9 | 22.6 | 23.2 |
| ≥10% | 13,568 | 101,114 | 13.4 | 13.2 | 13.6 |
| 5-10% | 9,583 | 101,114 | 9.5 | 9.3 | 9.7 |
| <5% | 25,943 | 101,114 | 25.7 | 25.4 | 25.9 |
| No wt | 52,020 | 101,114 | 51.4 | 51.1 | 51.8 |
| **12-Month Weight Outcome (Persistent at 12m, n = 133,436)** | | | | | |
| **Reached maintenance dose (≥1.7 mg; n = 70,463)** | | | | | |
| ≥5% | 32,460 | 70,463 | 46.1 | 45.7 | 46.4 |
| ≥10% | 24,574 | 70,463 | 34.9 | 34.5 | 35.2 |
| 5-10% | 7,886 | 70,463 | 11.2 | 11.0 | 11.4 |
| <5% | 6,709 | 70,463 | 9.5 | 9.3 | 9.7 |
| No wt | 31,294 | 70,463 | 44.4 | 44.0 | 44.8 |
| **Did NOT reach maintenance dose (n = 60,440)** | | | | | |
| ≥5% | 14,167 | 60,440 | 23.4 | 23.1 | 23.8 |
| ≥10% | 8,254 | 60,440 | 13.7 | 13.4 | 13.9 |
| 5-10% | 5,913 | 60,440 | 9.8 | 9.5 | 10.0 |
| <5% | 16,882 | 60,440 | 27.9 | 27.6 | 28.3 |
| No wt | 29,391 | 60,440 | 48.6 | 48.2 | 49.0 |
| **Dose Missing (n = 2,533)** | | | | | |
| ≥5% | 599 | 2,533 | 23.6 | 22.0 | 25.3 |
| ≥10% | 378 | 2,533 | 14.9 | 13.6 | 16.4 |
| 5-10% | 221 | 2,533 | 8.7 | 7.7 | 9.9 |
| <5% | 680 | 2,533 | 26.8 | 25.2 | 28.6 |
| No wt | 1,254 | 2,533 | 49.5 | 47.6 | 51.5 |

N denotes the number of patients achieving each weight-outcome category, and Denominator denotes the total number of patients in the corresponding care-pattern subgroup. PCT denotes the percentage of the denominator achieving each weight-loss category (≥5%, ≥10%, 5–10%, or <5% total body weight loss, or no weight recorded); 95% confidence intervals were calculated using the Wilson score method.

**Supplement Table 5. Weight-Loss Outcomes of Tirzepatide Initiators by Care Pattern.**

| | N | Denominator | PCT (%) | 95% CI Low | 95% CI High |
|---|---|---|---|---|---|
| **3-Month Weight Outcome (Non-persistent by 3m, n = 90,736)** | | | | | |
| ≥5% | 10,474 | 90,736 | 11.5 | 11.3 | 11.8 |
| ≥10% | 2,456 | 90,736 | 2.7 | 2.6 | 2.8 |
| 5-10% | 8,018 | 90,736 | 8.8 | 8.7 | 9.0 |
| <5% | 31,758 | 90,736 | 35.0 | 34.7 | 35.3 |
| No wt | 48,504 | 90,736 | 53.5 | 53.1 | 53.8 |
| **6-Month Weight Outcome (Non-persistent by 6m, n = 43,528)** | | | | | |
| ≥5% | 7,529 | 43,528 | 17.3 | 16.9 | 17.7 |
| ≥10% | 3,695 | 43,528 | 8.5 | 8.2 | 8.8 |
| 5-10% | 3,834 | 43,528 | 8.8 | 8.5 | 9.1 |
| <5% | 12,451 | 43,528 | 28.6 | 28.2 | 29.0 |
| No wt | 23,548 | 43,528 | 54.1 | 53.6 | 54.6 |
| **12-Month Weight Outcome (Non-persistent by 12m, n = 56,042)** | | | | | |
| ≥5% | 14,774 | 56,042 | 26.4 | 26.0 | 26.7 |
| ≥10% | 10,336 | 56,042 | 18.4 | 18.1 | 18.8 |
| 5-10% | 4,438 | 56,042 | 7.9 | 7.7 | 8.1 |
| <5% | 11,221 | 56,042 | 20.0 | 19.7 | 20.4 |
| No wt | 30,047 | 56,042 | 53.6 | 53.2 | 54.0 |
| **12-Month Weight Outcome (Persistent at 12m, n = 106,511)** | | | | | |
| **Reached maintenance dose (≥5.0 mg; n = 88,997)** | | | | | |
| ≥5% | 42,447 | 88,997 | 47.7 | 47.4 | 48.0 |
| ≥10% | 35,922 | 88,997 | 40.4 | 40.0 | 40.7 |
| 5-10% | 6,525 | 88,997 | 7.3 | 7.2 | 7.5 |
| <5% | 6,192 | 88,997 | 7.0 | 6.8 | 7.1 |
| No wt | 40,358 | 88,997 | 45.3 | 45.0 | 45.7 |
| **Did NOT reach maintenance dose (n = 16,585)** | | | | | |
| ≥5% | 2,841 | 16,585 | 17.1 | 16.6 | 17.7 |
| ≥10% | 1,666 | 16,585 | 10.0 | 9.6 | 10.5 |
| 5-10% | 1,175 | 16,585 | 7.1 | 6.7 | 7.5 |
| <5% | 5,383 | 16,585 | 32.5 | 31.7 | 33.2 |
| No wt | 8,361 | 16,585 | 50.4 | 49.7 | 51.2 |
| **Dose Missing (n = 929)** | | | | | |
| ≥5% | 284 | 929 | 30.6 | 27.7 | 33.6 |
| ≥10% | 214 | 929 | 23.0 | 20.4 | 25.9 |
| 5-10% | 70 | 929 | 7.5 | 6.0 | 9.4 |
| <5% | 196 | 929 | 21.1 | 18.6 | 23.8 |
| No wt | 449 | 929 | 48.3 | 45.1 | 51.5 |

N denotes the number of patients achieving each weight-outcome category, and Denominator denotes the total number of patients in the corresponding care-pattern subgroup. PCT denotes the percentage of the denominator achieving each weight-loss category (≥5%, ≥10%, 5–10%, or <5% total body weight loss, or no weight recorded); 95% confidence intervals were calculated using the Wilson score method.

**Supplement Table 6. Weight-Loss Outcomes At 6- and 12-Month of Semaglutide Initiators.**

| | **All Initiators** | | **Initiators with Weight Recorded** | | **Persistent Users with Weight Recorded** | | **Persistent at a maintenance dose with a recorded weight** | |
|---|---|---|---|---|---|---|---|---|
| | **≥5% TBWL** | **≥10% TBWL** | **≥5% TBWL** | **≥10% TBWL** | **≥5% TBWL** | **≥10% TBWL** | **≥5% TBWL** | **≥10% TBWL** |
| **12-Month Overall** | **25.6% (25.5-25.7)** | **16.4% (16.3-16.5)** | **51.9% (51.7-52.1)** | **33.2% (33.0-33.4)** | **66.1% (65.7-66.4)** | **46.4% (46.1-46.8)** | **82.9% (82.5-83.2)** | **62.7% (62.3-63.2)** |
| Sex | | | | | | | | |
| Female | 26.9% (26.7-27.0) | 17.9% (17.8-18.1) | 53.4% (53.2-53.7) | 35.7% (35.5-35.9) | 68.7% (68.3-69.1) | 50.7% (50.3-51.2) | 85.0% (84.6-85.4) | 67.4% (66.8-67.9) |
| Male | 22.2% (22.0-22.4) | 12.2% (12.0-12.4) | 47.6% (47.2-48.0) | 26.1% (25.7-26.4) | 58.7% (58.0-59.4) | 34.6% (33.9-35.3) | 76.8% (75.9-77.6) | 49.3% (48.4-50.3) |
| Race/Ethnicity | | | | | | | | |
| Hispanic | 22.9% (22.5-23.3) | 13.8% (13.4-14.1) | 49.9% (49.2-50.6) | 30.0% (29.3-30.7) | 61.9% (60.7-63.1) | 40.1% (38.9-41.3) | 80.5% (79.1-81.8) | 56.4% (54.7-58.1) |
| NH White | 27.6% (27.4-27.7) | 18.1% (18.0-18.3) | 54.0% (53.7-54.2) | 35.5% (35.3-35.8) | 69.1% (68.7-69.6) | 50.3% (49.9-50.8) | 84.5% (84.1-85.0) | 65.9% (65.3-66.5) |
| NH Black | 22.1% (21.8-22.5) | 12.5% (12.3-12.8) | 44.1% (43.6-44.7) | 25.0% (24.5-25.5) | 57.5% (56.6-58.5) | 36.1% (35.2-37.0) | 77.3% (76.2-78.4) | 53.4% (51.7-54.7) |
| NH Asian | 21.8% (20.8-22.8) | 13.0% (12.2-13.8) | 49.6% (47.7-51.4) | 29.5% (27.8-31.2) | 58.7% (55.6-61.6) | 36.4% (33.5-39.3) | 73.9% (70.0-77.4) | 49.3% (45.0-53.5) |
| SVI | | | | | | | | |
| Q1 | 27.8% (27.5-28.1) | 18.4% (18.1-18.7) | 56.5% (56.0-57.0) | 37.5% (37.0-37.9) | 69.9% (69.2-70.7) | 50.2% (49.4-51.0) | 84.9% (84.1-85.7) | 65.1% (64.0-66.1) |
| Q2 | 26.8% (26.6-27.1) | 17.6% (17.4-17.8) | 54.3% (53.8-54.7) | 35.6% (35.2-36.0) | 68.5% (67.8-69.2) | 49.6% (48.8-50.3) | 84.3% (83.6-85.0) | 65.2% (64.3-66.2) |
| Q3 | 25.3% (25.1-25.6) | 16.2% (15.9-16.4) | 51.0% (50.6-51.4) | 32.6% (32.2-32.9) | 65.8% (65.1-66.5) | 46.5% (45.8-47.3) | 82.4% (81.6-83.1) | 62.7% (61.7-63.6) |
| Q4 | 23.6% (23.4-23.8) | 14.4% (14.2-14.6) | 48.0% (47.6-48.3) | 29.2% (28.9-29.6) | 61.6% (60.9-62.2) | 41.3% (40.6-41.9) | 80.5% (79.7-81.2) | 58.7% (57.8-59.7) |
| Insurance Type | | | | | | | | |
| Commercial | 25.5% (25.4-25.7) | 16.6% (16.5-16.8) | 53.8% (53.5-54.0) | 35.0% (34.8-35.3) | 67.0% (66.5-67.4) | 47.6% (47.1-48.0) | 83.3% (82.9-83.8) | 63.2% (62.7-63.8) |
| Medicaid/ Replacement | 24.7% (24.3-25.1) | 15.9% (15.6-16.3) | 51.2% (50.6-51.9) | 33.0% (32.4-33.6) | 64.9% (63.8-65.9) | 45.6% (44.5-46.8) | 80.8% (79.6-82.0) | 61.4% (59.9-62.8) |
| Medicare/ Advantage | 27.7% (27.4-28.1) | 16.0% (15.7-16.3) | 45.2% (44.7-45.7) | 26.1% (25.6-26.5) | 63.0% (61.9-64.0) | 41.7% (40.7-42.8) | 82.2% (80.8-83.5) | 61.3% (59.6-62.9) |
| Self-pay | 18.7% (17.0-20.5) | 10.6% (9.3-12.0) | 48.9% (45.3-52.5) | 27.7% (24.6-31.1) | 56.7% (50.8-62.5) | 31.3% (26.1-37.0) | 72.8% (64.0-80.1) | 50.9% (41.8-59.9) |
| **6-Month Overall** | **21.7% (21.6-21.8)** | **10.2% (10.1-10.3)** | **44.8% (44.6-45.1)** | **21.1% (20.9-21.3)** | **53.3% (53.0-53.6)** | **26.2% (25.9-26.4)** | **74.1% (73.7-74.5)** | **41.3% (40.8-41.8)** |

| | | | | | | | | |
|---|---|---|---|---|---|---|---|---|
| Sex | | | | | | | | |
| Female | 22.9% | 11.3% | 46.2% | 22.7% | 55.5% | 28.7% | 76.0% | 44.7% |
| | (22.7-23.0) | (11.2-11.4) | (46.0-46.5) | (22.5-23.0) | (55.1-55.8) | (28.4-29.0) | (75.5-76.5) | (44.2-45.3) |
| Male | 18.6% | 7.4% | 40.9% | 16.3% | 47.3% | 19.1% | 68.6% | 31.4% |
| | (18.3-18.8) | (7.3-7.5) | (40.4-41.3) | (16.0-16.6) | (46.8-47.9) | (18.7-19.5) | (67.8-69.5) | (30.6-32.3) |
| Race/Ethnicity | | | | | | | | |
| Hispanic | 20.5% | 8.6% | 43.3% | 18.2% | 50.6% | 21.9% | 70.8% | 36.4% |
| | (20.2-21.0) | (8.4-8.9) | (42.6-44.0) | (17.7-18.8) | (49.6-51.5) | (21.2-22.7) | (69.4-72.3) | (34.9-38.0) |
| NH White | 23.1% | 11.4% | 46.7% | 22.9% | 55.8% | 28.7% | 75.7% | 44.1% |
| | (23.0-23.3) | (11.2-11.5) | (46.4-47.0) | (22.7-23.2) | (55.4-56.1) | (28.4-29.1) | (75.2-76.2) | (43.5-44.7) |
| NH Black | 18.5% | 7.4% | 37.0% | 14.7% | 44.8% | 18.6% | 68.4% | 32.5% |
| | (18.2-18.8) | (7.2-7.6) | (36.5-37.5) | (14.3-15.1) | (44.1-45.5) | (18.0-19.1) | (67.2-69.5) | (31.4-33.7) |
| NH Asian | 18.4% | 7.5% | 43.2% | 17.6% | 49.6% | 19.7% | 66.1% | 29.3% |
| | (17.5-19.4) | (6.9-8.2) | (41.3-45.0) | (16.2-19.0) | (47.2-52.0) | (17.9-21.7) | (62.3-69.8) | (25.9-33.0) |
| SVI | | | | | | | | |
| Q1 | 22.9% | 11.3% | 49.5% | 24.4% | 58.2% | 29.8% | 76.9% | 43.8% |
| | (22.6-23.1) | (11.1-11.5) | (49.0-50.0) | (24.0-24.8) | (57.6-58.9) | (29.2-30.4) | (76.0-77.7) | (42.8-44.8) |
| Q2 | 22.5% | 11.0% | 47.1% | 23.0% | 55.9% | 28.4% | 75.1% | 43.0% |
| | (22.3-22.8) | (10.8-11.2) | (46.7-47.5) | (22.6-23.4) | (55.3-56.4) | (27.9-28.9) | (74.3-75.9) | (42.1-44.0) |
| Q3 | 21.6% | 10.2% | 44.1% | 20.8% | 52.4% | 25.9% | 73.7% | 41.6% |
| | (21.4-21.8) | (10.0-10.4) | (43.7-44.5) | (20.4-21.1) | (51.8-52.9) | (25.4-26.4) | (72.8-74.5) | (40.7-42.6) |
| Q4 | 20.5% | 9.0% | 41.0% | 18.0% | 49.2% | 22.5% | 71.7% | 37.7% |
| | (20.3-20.7) | (8.8-9.1) | (40.7-41.4) | (17.7-18.3) | (48.7-49.7) | (22.1-22.9) | (70.9-72.5) | (36.9-38.6) |
| Insurance Type | | | | | | | | |
| Commercial | 21.8% | 10.6% | 47.4% | 22.9% | 55.6% | 27.9% | 75.1% | 42.2% |
| | (21.7-22.0) | (10.4-10.7) | (47.2-47.7) | (22.7-23.2) | (55.2-55.9) | (27.6-28.2) | (74.6-75.6) | (41.7-42.8) |
| Medicaid/ | 22.8% | 10.6% | 44.3% | 20.7% | 53.0% | 25.6% | 72.3% | 39.9% |
| Replacement | (22.5-23.2) | (10.4-10.9) | (43.7-45.0) | (20.2-21.2) | (52.1-53.8) | (24.8-26.3) | (71.1-73.5) | (38.6-41.2) |
| Medicare/ | 20.9% | 8.4% | 35.2% | 14.2% | 43.7% | 18.7% | 69.7% | 35.6% |
| Advantage | (20.5-21.2) | (8.2-8.7) | (34.6-35.7) | (13.8-14.6) | (42.9-44.5) | (18.0-19.3) | (68.1-71.2) | (34.0-37.3) |
| Self-pay | 19.2% | 8.8% | 45.7% | 21.0% | 53.1% | 24.7% | 67.2% | 37.1% |
| | (17.5-21.0) | (7.6-10.2) | (42.2-49.1) | (18.3-23.9) | (48.6-57.5) | (21.1-28.7) | (58.3-75.1) | (28.8-46.1) |

Values are percentages (95% confidence interval) of patients achieving ≥5% or ≥10% total body weight loss (TBWL) at 6 and 12 months, among all semaglutide initiators, semaglutide initiators with a recorded weight, prescribing persistent users with a recorded weight, and prescribing persistent users at an initial FDA-labeled maintenance dose with a recorded weight, overall and stratified by sex, race/ethnicity, Social Vulnerability Index (SVI) quartile, and insurance type.

**Supplement Table 7. Weight-Loss Outcomes At 6- and 12-Month of Tirzepatide Initiators.**

| | All Initiators | | Initiators with Weight Recorded | | Persistent Users with Weight Recorded | | Persistent at a maintenance dose with a recorded weight | |
|---|---|---|---|---|---|---|---|---|
| | ≥5% TBWL | ≥10% TBWL | ≥5% TBWL | ≥10% TBWL | ≥5% TBWL | ≥10% TBWL | ≥5% TBWL | ≥10% TBWL |
| **12-Month Overall** | **30.4% (30.2-30.6)** | **22.5% (22.3-22.6)** | **61.9% (61.6-62.1)** | **45.8% (45.5-46.0)** | **79.5% (79.1-79.8)** | **65.9% (65.5-66.3)** | **87.3% (87.0-87.6)** | **73.9% (73.5-74.2)** |
| Sex | | | | | | | | |
| Female | 31.7% (31.5-31.9) | 24.2% (24.0-24.4) | 63.2% (62.9-63.5) | 48.1% (47.9-48.4) | 81.6% (81.2-82.0) | 69.7% (69.2-70.1) | 88.9% (88.6-89.2) | 77.4% (76.9-77.8) |
| Male | 27.1% (26.8-27.4) | 18.2% (17.9-18.5) | 58.3% (57.8-58.8) | 39.2% (38.7-39.7) | 73.9% (73.2-74.6) | 56.2% (55.4-57.0) | 82.9% (82.2-83.5) | 64.5% (63.6-65.3) |
| Race/Ethnicity | | | | | | | | |
| Hispanic | 27.9% (27.3-28.6) | 20.0% (19.5-20.6) | 59.5% (58.5-60.4) | 42.6% (41.6-43.6) | 76.4% (75.0-77.8) | 62.3% (60.7-63.8) | 84.4% (83.1-85.6) | 70.4% (68.8-72.0) |
| NH White | 32.2% (32.0-32.4) | 24.3% (24.1-24.5) | 63.8% (63.5-64.1) | 48.2% (47.8-48.5) | 81.5% (81.1-81.9) | 68.6% (68.1-69.1) | 88.6% (88.3-89.0) | 76.0% (75.6-76.5) |
| NH Black | 26.1% (25.7-26.6) | 17.3% (16.9-17.7) | 52.5% (51.8-53.3) | 34.8% (34.2-35.5) | 71.6% (70.5-72.7) | 54.7% (53.5-56.0) | 81.5% (80.4-82.5) | 64.0% (62.7-65.3) |
| NH Asian | 26.7% (25.4-28.1) | 19.2% (18.1-20.4) | 62.6% (60.4-64.8) | 45.1% (42.8-47.4) | 75.9% (73.0-78.7) | 60.6% (57.3-63.8) | 84.5% (81.6-86.9) | 68.5% (65.0-71.8) |
| SVI | | | | | | | | |
| Q1 | 32.7% (32.3-33.0) | 25.0% (24.7-25.4) | 67.2% (66.7-67.7) | 51.5% (51.0-52.1) | 82.2% (81.5-82.8) | 69.2% (68.5-70.0) | 88.9% (88.4-89.5) | 76.2% (75.4-76.9) |
| Q2 | 31.5% (31.2-31.9) | 23.6% (23.3-23.9) | 64.0% (63.5-64.5) | 47.9% (47.4-48.5) | 81.1% (80.5-81.7) | 67.8% (67.1-68.5) | 88.2% (87.6-88.7) | 75.1% (74.4-75.8) |
| Q3 | 29.9% (29.6-30.2) | 21.9% (21.6-22.2) | 60.5% (60.0-61.0) | 44.3% (43.7-44.8) | 78.6% (77.9-79.3) | 64.8% (64.0-65.6) | 87.0% (86.3-87.6) | 73.2% (72.3-73.9) |
| Q4 | 28.0% (27.7-28.3) | 19.9% (19.6-20.2) | 56.6% (56.1-57.1) | 40.3% (39.8-40.8) | 75.7% (75.0-76.4) | 61.6% (60.8-62.4) | 84.7% (84.1-85.4) | 70.6% (69.7-71.4) |
| Insurance Type | | | | | | | | |
| Commercial | 30.8% (30.6-31.0) | 23.2% (23.0-23.3) | 63.9% (63.6-64.2) | 48.1% (47.8-48.4) | 80.2% (79.8-80.6) | 66.8% (66.4-67.3) | 87.5% (87.2-87.9) | 74.3% (73.8-74.7) |
| Medicaid/ Replacement | 27.3% (26.6-27.9) | 19.5% (18.9-20.1) | 56.0% (54.9-57.1) | 40.0% (39.0-41.1) | 76.3% (74.6-77.9) | 63.8% (61.9-65.6) | 85.3% (83.7-86.7) | 73.1% (71.1-74.9) |
| Medicare/ Advantage | 31.5% (30.9-32.1) | 20.2% (19.7-20.7) | 51.7% (50.8-52.5) | 33.1% (32.3-33.8) | 73.9% (72.5-75.3) | 57.4% (55.8-58.9) | 85.5% (84.1-86.7) | 68.5% (66.8-70.2) |
| Self-pay | 24.3% (21.6-27.3) | 16.5% (14.2-19.1) | 63.0% (57.7-68.1) | 42.7% (37.5-48.1) | 76.2% (68.7-82.4) | 55.8% (47.7-63.6) | 84.8% (77.5-90.0) | 63.2% (54.5-71.1) |
| **6-Month Overall** | **26.9% (26.7-27.0)** | **16.3% (16.1-16.4)** | **56.0% (55.8-56.3)** | **33.9% (33.7-34.2)** | **68.9% (68.5-69.2)** | **45.7% (45.4-46.0)** | **80.9% (80.6-81.2)** | **55.6% (55.2-56.0)** |

| | | | | | | | | |
|---|---|---|---|---|---|---|---|---|
| Sex | | | | | | | | |
| Female | 28.0% (27.8-28.2) | 17.4% (17.2-17.6) | 57.0% (56.7-57.3) | 35.4% (35.1-35.7) | 70.5% (70.1-70.8) | 48.1% (47.7-48.5) | 82.2% (81.9-82.6) | 58.1% (57.7-58.6) |
| Male | 24.0% (23.7-24.3) | 13.4% (13.1-13.6) | 53.3% (52.8-53.8) | 29.7% (29.3-30.2) | 64.6% (64.0-65.2) | 39.2% (38.6-39.8) | 77.2% (76.6-77.8) | 48.7% (48.0-49.5) |
| Race/Ethnicity | | | | | | | | |
| Hispanic | 25.7% (25.1-26.3) | 14.5% (14.0-15.0) | 53.7% (52.7-54.7) | 30.3% (29.4-31.2) | 66.8% (65.6-68.1) | 41.5% (40.2-42.8) | 79.0% (77.8-80.2) | 51.1% (49.6-52.6) |
| NH White | 28.3% (28.1-28.5) | 17.6% (17.5-17.8) | 57.6% (57.3-58.0) | 36.0% (35.6-36.3) | 70.6% (70.2-71.0) | 48.1% (47.6-48.5) | 82.3% (81.9-82.6) | 58.0% (57.5-58.5) |
| NH Black | 22.6% (22.2-23.1) | 11.7% (11.3-12.0) | 46.2% (45.4-46.9) | 23.8% (23.2-24.4) | 59.1% (58.1-60.1) | 34.2% (33.2-35.1) | 72.6% (71.5-73.6) | 43.8% (42.6-45.0) |
| NH Asian | 24.9% (23.6-26.2) | 14.2% (13.2-15.3) | 58.5% (56.2-60.8) | 33.4% (31.3-35.6) | 69.3% (66.6-71.9) | 43.7% (40.9-46.6) | 79.8% (77.0-82.2) | 52.0% (48.8-55.3) |
| SVI | | | | | | | | |
| Q1 | 28.4% (28.1-28.7) | 18.0% (17.7-18.2) | 61.6% (61.1-62.2) | 39.0% (38.4-39.5) | 73.5% (72.9-74.2) | 50.6% (49.9-51.3) | 83.1% (82.5-83.7) | 58.7% (57.9-59.5) |
| Q2 | 27.8% (27.5-28.1) | 17.3% (17.0-17.5) | 58.1% (57.6-58.7) | 36.1% (35.6-36.6) | 70.7% (70.0-71.3) | 47.8% (47.1-48.5) | 81.9% (81.3-82.5) | 57.3% (56.5-58.0) |
| Q3 | 26.6% (26.3-26.9) | 15.8% (15.5-16.0) | 54.7% (54.2-55.2) | 32.5% (32.0-33.0) | 67.7% (67.1-68.4) | 44.4% (43.7-45.1) | 80.7% (80.0-81.3) | 54.9% (54.1-55.6) |
| Q4 | 25.0% (24.7-25.4) | 14.4% (14.1-14.6) | 50.7% (50.2-51.2) | 29.1% (28.7-29.6) | 63.9% (63.3-64.6) | 40.4% (39.8-41.1) | 77.9% (77.2-78.6) | 51.4% (50.6-52.2) |
| Insurance Type | | | | | | | | |
| Commercial | 27.4% (27.2-27.6) | 17.0% (16.8-17.1) | 58.7% (58.4-59.0) | 36.4% (36.1-36.7) | 70.9% (70.5-71.2) | 47.7% (47.3-48.0) | 81.7% (81.3-82.0) | 56.6% (56.2-57.1) |
| Medicaid/ Replacement | 25.4% (24.7-26.1) | 14.3% (13.7-14.8) | 49.4% (48.3-50.5) | 27.7% (26.8-28.7) | 63.7% (62.2-65.1) | 40.8% (39.3-42.3) | 77.3% (75.7-78.7) | 51.4% (49.6-53.2) |
| Medicare/ Advantage | 24.5% (23.9-25.0) | 12.2% (11.8-12.6) | 41.8% (41.0-42.7) | 20.8% (20.2-21.5) | 54.8% (53.5-56.0) | 31.4% (30.3-32.6) | 74.4% (73.0-75.8) | 45.9% (44.3-47.6) |
| Self-pay | 21.1% (18.5-23.9) | 13.0% (10.9-15.4) | 52.3% (47.1-57.5) | 32.3% (27.5-37.4) | 61.7% (54.8-68.3) | 40.3% (33.7-47.3) | 73.8% (66.2-80.2) | 50.3% (42.4-58.3) |

Values are percentages (95% confidence interval) of patients achieving ≥5% or ≥10% total body weight loss (TBWL) at 6 and 12 months, among all tirzepatide initiators, tirzepatide initiators with a recorded weight, persistent users with a recorded weight, and persistent users at an initial FDA-labeled maintenance dose with a recorded weight, overall and stratified by sex, race/ethnicity, Social Vulnerability Index (SVI) quartile, and insurance type.

**Supplement Table 8. Weight-Loss Outcomes At 6- and 12-Month of WEGOVY Initiators.**

| | All Initiators | | Initiators with Weight Recorded | | Persistent Users with Weight Recorded | | Persistent at a maintenance dose with a recorded weight | |
|---|---|---|---|---|---|---|---|---|
| | ≥5% TBWL | ≥10% TBWL | ≥5% TBWL | ≥10% TBWL | ≥5% TBWL | ≥10% TBWL | ≥5% TBWL | ≥10% TBWL |
| **12-Month Overall** | 24.6% (24.4-24.8) | 15.6% (15.4-15.8) | 49.8% (49.5-50.1) | 31.6% (31.3-31.8) | 63.4% (62.9-64.0) | 44.6% (44.1-45.2) | 82.9% (82.3-83.5) | 63.2% (62.4-64.0) |
| Sex | | | | | | | | |
| Female | 25.8% (25.6-26.1) | 17.1% (16.9-17.3) | 51.2% (50.9-51.6) | 33.9% (33.5-34.2) | 66.1% (65.5-66.8) | 48.8% (48.1-49.5) | 85.0% (84.3-85.6) | 67.7% (66.8-68.5) |
| Male | 21.2% (20.8-21.5) | 11.5% (11.3-11.8) | 45.3% (44.7-46.0) | 24.7% (24.2-25.2) | 55.8% (54.6-56.9) | 32.8% (31.8-33.9) | 76.6% (75.2-77.9) | 49.8% (48.2-51.4) |
| Race/Ethnicity | | | | | | | | |
| Hispanic | 22.2% (21.6-22.9) | 13.5% (13.0-14.0) | 48.6% (47.5-49.7) | 29.5% (28.5-30.4) | 61.1% (59.1-63.0) | 40.1% (38.2-42.1) | 81.5% (79.3-83.6) | 58.6% (55.8-61.3) |
| NH White | 26.5% (26.2-26.7) | 17.2% (17.0-17.5) | 51.6% (51.2-52.0) | 33.6% (33.2-34.0) | 66.4% (65.7-67.2) | 48.4% (47.6-49.2) | 84.5% (83.7-85.3) | 66.5% (65.5-67.5) |
| NH Black | 21.6% (21.2-22.1) | 12.3% (12.0-12.7) | 42.8% (42.1-43.6) | 24.4% (23.8-25.1) | 56.0% (54.6-57.4) | 36.1% (34.7-37.4) | 78.2% (76.4-79.8) | 55.2% (53.2-57.2) |
| NH Asian | 21.9% (20.4-23.4) | 13.5% (12.4-14.8) | 49.7% (47.0-52.4) | 30.8% (28.3-33.3) | 57.3% (52.6-61.8) | 36.8% (32.4-41.4) | 72.6% (66.7-77.9) | 51.9% (45.6-58.2) |
| SVI | | | | | | | | |
| Q1 | 26.8% (26.4-27.3) | 17.4% (17.1-17.8) | 54.1% (53.4-54.9) | 35.2% (34.5-35.9) | 67.4% (66.1-68.7) | 48.2% (46.8-49.5) | 84.9% (83.6-86.2) | 65.4% (63.6-67.1) |
| Q2 | 25.8% (25.4-26.1) | 16.8% (16.5-17.2) | 51.8% (51.1-52.4) | 33.8% (33.2-34.4) | 65.5% (64.3-66.6) | 47.7% (46.5-48.9) | 83.7% (82.5-84.8) | 65.8% (64.3-67.3) |
| Q3 | 24.2% (23.9-24.6) | 15.4% (15.1-15.7) | 48.8% (48.2-49.4) | 31.0% (30.4-31.5) | 63.3% (62.2-64.4) | 45.0% (43.8-46.1) | 82.5% (81.3-83.7) | 63.7% (62.2-65.2) |
| Q4 | 22.9% (22.6-23.2) | 13.9% (13.6-14.2) | 46.7% (46.2-47.2) | 28.3% (27.9-28.8) | 59.7% (58.7-60.7) | 40.1% (39.1-41.1) | 81.3% (80.1-82.4) | 59.4% (58.0-60.8) |
| Insurance Type | | | | | | | | |
| Commercial | 24.5% (24.3-24.8) | 15.9% (15.7-16.0) | 51.5% (51.1-51.8) | 33.2% (32.9-33.6) | 64.2% (63.5-64.8) | 45.6% (44.9-46.3) | 83.0% (82.3-83.7) | 63.5% (62.5-64.4) |
| Medicaid/ Replacement | 23.6% (23.0-24.2) | 14.9% (14.4-15.4) | 49.2% (48.2-50.2) | 31.2% (30.3-32.1) | 61.4% (59.5-63.3) | 42.7% (40.8-44.6) | 80.1% (77.9-82.1) | 60.6% (58.0-63.1) |
| Medicare/ Advantage | 26.0% (25.6-26.6) | 14.6% (14.2-15.1) | 42.1% (41.3-42.9) | 23.6% (23.0-24.3) | 58.2% (56.4-60.1) | 37.6% (35.8-39.4) | 84.1% (81.7-86.2) | 63.0% (60.0-65.9) |
| Self-pay | 18.7% (15.6-22.2) | 10.4% (8.1-13.2) | 51.5% (44.6-58.4) | 28.6% (22.7-35.3) | 60.3% (48.0-71.5) | 34.9% (24.3-47.2) | 78.1% (61.2-89.0) | 50.0% (33.6-66.4) |
| **6-Month Overall** | 20.6% (20.5-20.8) | 9.7% (9.6-9.9) | 42.4% (42.1-42.7) | 20.0% (19.8-20.3) | 50.4% (49.9-50.8) | 24.9% (24.5-25.3) | 73.7% (73.1-74.3) | 41.2% (40.5-41.9) |

| | | | | | | | | |
|---|---|---|---|---|---|---|---|---|
| Sex | | | | | | | | |
| Female | 21.7% (21.5-21.9) | 10.7% (10.6-10.9) | 43.6% (43.3-44.0) | 21.5% (21.2-21.8) | 52.3% (51.8-52.9) | 27.3% (26.8-27.8) | 75.6% (74.8-76.3) | 44.6% (43.7-45.4) |
| Male | 17.7% (17.4-18.0) | 7.1% (6.9-7.3) | 38.9% (38.3-39.5) | 15.5% (15.1-16.0) | 44.7% (43.8-45.5) | 17.9% (17.3-18.6) | 68.2% (66.9-69.6) | 31.2% (29.8-32.5) |
| Race/Ethnicity | | | | | | | | |
| Hispanic | 19.8% (19.2-20.4) | 8.5% (8.1-8.9) | 41.4% (40.4-42.5) | 17.9% (17.1-18.7) | 49.2% (47.7-50.7) | 21.6% (20.4-22.8) | 71.3% (69.0-73.4) | 36.6% (34.3-38.9) |
| NH White | 22.0% (21.8-22.3) | 10.8% (10.6-11.0) | 44.0% (43.6-44.4) | 21.6% (21.2-21.9) | 52.4% (51.8-52.9) | 27.0% (26.5-27.6) | 75.4% (74.5-76.2) | 43.9% (43.0-44.9) |
| NH Black | 17.9% (17.5-18.3) | 7.3% (7.0-7.6) | 35.9% (35.2-36.7) | 14.7% (14.1-15.2) | 43.3% (42.2-44.4) | 18.7% (17.9-19.6) | 68.6% (66.9-70.2) | 33.7% (32.0-35.4) |
| NH Asian | 18.8% (17.4-20.2) | 7.6% (6.7-8.6) | 43.4% (40.7-46.1) | 17.5% (15.5-19.6) | 48.6% (44.9-52.3) | 19.4% (16.7-22.5) | 68.1% (62.2-73.4) | 29.6% (24.4-35.4) |
| SVI | | | | | | | | |
| Q1 | 21.9% (21.5-22.3) | 10.7% (10.3-11.0) | 46.7% (46.0-47.5) | 22.7% (22.1-23.4) | 55.0% (53.9-56.0) | 28.0% (27.1-29.0) | 76.3% (74.9-77.7) | 42.9% (41.3-44.6) |
| Q2 | 21.3% (20.9-21.6) | 10.4% (10.1-10.7) | 44.5% (43.8-45.1) | 21.7% (21.2-22.2) | 53.0% (52.1-53.9) | 27.1% (26.3-27.9) | 74.6% (73.3-75.8) | 43.1% (41.7-44.6) |
| Q3 | 20.6% (20.2-20.9) | 9.8% (9.6-10.1) | 41.7% (41.1-42.3) | 20.0% (19.5-20.4) | 49.3% (48.4-50.2) | 24.7% (24.0-25.5) | 73.5% (72.2-74.8) | 42.4% (41.0-43.8) |
| Q4 | 19.6% (19.3-19.9) | 8.7% (8.5-8.9) | 39.3% (38.8-39.8) | 17.5% (17.1-17.9) | 46.9% (46.2-47.7) | 21.8% (21.2-22.5) | 71.7% (70.5-72.8) | 37.8% (36.6-39.1) |
| Insurance Type | | | | | | | | |
| Commercial | 20.8% (20.6-21.0) | 10.0% (9.9-10.2) | 45.0% (44.6-45.4) | 21.7% (21.4-22.0) | 52.6% (52.1-53.2) | 26.5% (26.0-26.9) | 74.2% (73.4-74.9) | 41.6% (40.8-42.5) |
| Medicaid/ Replacement | 21.4% (20.9-22.0) | 9.8% (9.4-10.3) | 41.4% (40.4-42.3) | 19.0% (18.3-19.8) | 49.9% (48.6-51.3) | 23.6% (22.4-24.7) | 72.2% (70.2-74.1) | 38.8% (36.7-40.9) |
| Medicare/ Advantage | 19.0% (18.5-19.5) | 7.7% (7.3-8.0) | 31.5% (30.8-32.3) | 12.7% (12.2-13.3) | 37.9% (36.7-39.2) | 16.4% (15.5-17.4) | 70.1% (67.3-72.7) | 37.1% (34.3-40.0) |
| Self-pay | 16.1% (13.3-19.4) | 7.8% (5.8-10.3) | 42.4% (35.9-49.3) | 20.5% (15.5-26.5) | 52.2% (43.1-61.2) | 23.9% (17.0-32.5) | 75.0% (56.6-87.3) | 32.1% (17.9-50.7) |

Values are percentages (95% confidence interval) of patients achieving ≥5% or ≥10% total body weight loss (TBWL) at 6 and 12 months, among all WEGOVY initiators, WEGOVY initiators with a recorded weight, persistent users with a recorded weight, and persistent users at an initial FDA-labeled maintenance dose with a recorded weight, overall and stratified by sex, race/ethnicity, Social Vulnerability Index (SVI) quartile, and insurance type.

**Supplement Table 9. Weight-Loss Outcomes At 6- and 12-Month of ZEPBOUND Initiators.**

| | All Initiators | | Initiators with Weight Recorded | | Persistent Users with Weight Recorded | | Persistent at a maintenance dose with a recorded weight | |
|---|---|---|---|---|---|---|---|---|
| | ≥5% TBWL | ≥10% TBWL | ≥5% TBWL | ≥10% TBWL | ≥5% TBWL | ≥10% TBWL | ≥5% TBWL | ≥10% TBWL |
| **12-Month Overall** | 28.6% (28.4-28.9) | 20.7% (20.4-20.9) | 58.6% (58.2-59.0) | 42.3% (41.9-42.6) | 77.2% (76.7-77.8) | 63.7% (63.0-64.3) | 86.8% (86.4-87.3) | 73.5% (72.9-74.1) |
| Sex | | | | | | | | |
| Female | 29.9% (29.6-30.2) | 22.2% (22.0-22.5) | 59.9% (59.5-60.3) | 44.5% (44.1-44.9) | 79.6% (79.0-80.2) | 67.5% (66.7-68.1) | 88.5% (88.0-89.1) | 76.9% (76.2-77.6) |
| Male | 25.4% (24.9-25.8) | 16.5% (16.2-16.9) | 54.9% (54.2-55.7) | 35.8% (35.1-36.5) | 70.9% (69.8-72.0) | 53.6% (52.4-54.8) | 82.1% (81.0-83.1) | 64.0% (62.6-65.3) |
| Race/Ethnicity | | | | | | | | |
| Hispanic | 27.2% (26.3-28.0) | 19.3% (18.6-20.1) | 57.2% (55.8-58.5) | 40.7% (39.4-42.0) | 75.4% (73.4-77.4) | 61.4% (59.2-63.7) | 85.1% (83.2-86.8) | 71.3% (68.9-73.5) |
| NH White | 30.5% (30.2-30.8) | 22.4% (22.2-22.7) | 60.4% (60.0-60.9) | 44.5% (44.0-44.9) | 79.6% (78.9-80.2) | 66.8% (66.0-67.6) | 88.5% (87.9-89.1) | 76.1% (75.3-76.9) |
| NH Black | 24.6% (24.0-25.2) | 16.0% (15.4-16.5) | 50.2% (49.2-51.2) | 32.5% (31.6-33.5) | 69.8% (68.1-71.4) | 52.7% (50.8-54.5) | 81.5% (79.9-83.1) | 63.7% (61.7-65.7) |
| NH Asian | 25.8% (24.0-27.7) | 17.4% (15.9-19.0) | 59.2% (56.1-62.3) | 39.9% (36.8-43.0) | 72.6% (68.0-76.8) | 55.3% (50.3-60.2) | 83.2% (78.6-86.9) | 64.4% (59.0-69.5) |
| SVI | | | | | | | | |
| Q1 | 30.9% (30.4-31.5) | 23.1% (22.6-23.6) | 64.0% (63.2-64.8) | 47.8% (47.0-48.7) | 80.1% (79.0-81.2) | 67.1% (65.8-68.4) | 88.3% (87.4-89.3) | 75.8% (74.5-77.1) |
| Q2 | 29.7% (29.2-30.2) | 21.7% (21.3-22.1) | 60.7% (59.9-61.4) | 44.3% (43.5-45.0) | 79.2% (78.2-80.2) | 65.9% (64.7-67.0) | 88.1% (87.2-89.0) | 75.0% (73.7-76.1) |
| Q3 | 28.3% (27.9-28.8) | 20.3% (19.9-20.7) | 57.6% (56.9-58.3) | 41.3% (40.5-42.0) | 76.2% (75.1-77.3) | 62.6% (61.3-63.8) | 86.5% (85.4-87.4) | 73.1% (71.8-74.3) |
| Q4 | 26.5% (26.1-27.0) | 18.4% (18.1-18.8) | 54.0% (53.3-54.7) | 37.5% (36.8-38.2) | 73.9% (72.8-75.0) | 59.6% (58.4-60.9) | 84.6% (83.6-85.6) | 70.3% (69.0-71.6) |
| Insurance Type | | | | | | | | |
| Commercial | 28.9% (28.6-29.2) | 21.2% (20.9-21.4) | 60.5% (60.0-60.9) | 44.3% (43.9-44.7) | 78.0% (77.4-78.6) | 64.6% (63.9-65.2) | 87.0% (86.5-87.5) | 73.6% (72.9-74.3) |
| Medicaid/ Replacement | 26.1% (25.1-27.1) | 17.9% (17.0-18.8) | 53.9% (52.2-55.5) | 36.9% (35.3-38.5) | 74.3% (71.4-76.9) | 62.0% (58.9-65.0) | 84.4% (81.6-86.8) | 73.3% (70.1-76.3) |
| Medicare/ Advantage | 29.5% (28.6-30.4) | 17.9% (17.2-18.7) | 47.6% (46.4-48.9) | 29.0% (27.9-30.1) | 68.2% (65.7-70.7) | 51.6% (48.9-54.3) | 84.4% (81.8-86.7) | 68.4% (65.2-71.5) |
| Self-pay | 18.9% (15.0-23.6) | 12.4% (9.3-16.5) | 53.5% (44.4-62.4) | 35.1% (26.9-44.2) | 65.2% (50.8-77.3) | 45.7% (32.2-59.8) | 77.1% (61.0-87.9) | 57.1% (40.9-72.0) |
| **6-Month Overall** | 25.4% (25.1-25.6) | 15.1% (14.9-15.3) | 53.1% (52.7-53.5) | 31.6% (31.2-31.9) | 66.6% (66.1-67.1) | 44.2% (43.7-44.8) | 80.7% (80.2-81.2) | 55.9% (55.3-56.5) |

| | | | | | | | | |
|---|---|---|---|---|---|---|---|---|
| Sex | | | | | | | | |
| Female | 26.4% (26.1-26.7) | 16.1% (15.9-16.3) | 54.0% (53.5-54.4) | 32.9% (32.5-33.3) | 68.1% (67.5-68.7) | 46.6% (46.0-47.2) | 82.1% (81.5-82.6) | 58.5% (57.8-59.2) |
| Male | 22.6% (22.2-23.0) | 12.4% (12.1-12.7) | 50.6% (49.9-51.4) | 27.7% (27.1-28.4) | 62.3% (61.3-63.3) | 37.8% (36.8-38.8) | 76.9% (75.8-77.9) | 48.7% (47.5-49.9) |
| Race/Ethnicity | | | | | | | | |
| Hispanic | 24.6% (23.8-25.4) | 14.0% (13.4-14.7) | 51.6% (50.3-53.0) | 29.5% (28.2-30.7) | 65.7% (63.9-67.4) | 41.4% (39.6-43.3) | 79.4% (77.6-81.1) | 52.7% (50.5-54.9) |
| NH White | 27.0% (26.7-27.3) | 16.5% (16.3-16.8) | 54.9% (54.4-55.4) | 33.7% (33.2-34.1) | 68.4% (67.8-69.0) | 46.9% (46.2-47.6) | 82.3% (81.7-82.9) | 58.7% (58.0-59.5) |
| NH Black | 21.4% (20.9-22.0) | 10.9% (10.5-11.3) | 44.4% (43.4-45.5) | 22.6% (21.7-23.5) | 58.3% (56.9-59.8) | 33.7% (32.3-35.1) | 74.0% (72.4-75.5) | 44.7% (43.0-46.5) |
| NH Asian | 22.9% (21.2-24.7) | 12.1% (10.8-13.5) | 54.4% (51.2-57.6) | 28.7% (25.9-31.7) | 65.6% (61.4-69.5) | 39.1% (35.1-43.4) | 80.1% (75.9-83.8) | 49.0% (44.1-53.9) |
| SVI | | | | | | | | |
| Q1 | 27.1% (26.6-27.6) | 16.9% (16.5-17.3) | 59.0% (58.1-59.8) | 36.7% (35.9-37.5) | 71.7% (70.7-72.7) | 49.2% (48.1-50.4) | 83.2% (82.2-84.1) | 59.0% (57.7-60.3) |
| Q2 | 26.4% (25.9-26.9) | 16.1% (15.7-16.5) | 55.3% (54.5-56.0) | 33.8% (33.1-34.5) | 68.8% (67.8-69.7) | 47.0% (45.9-48.0) | 81.9% (81.0-82.8) | 58.1% (56.9-59.3) |
| Q3 | 25.1% (24.6-25.5) | 14.5% (14.2-14.9) | 52.0% (51.2-52.7) | 30.1% (29.4-30.8) | 65.3% (64.3-66.3) | 42.7% (41.6-43.7) | 80.4% (79.4-81.4) | 55.0% (53.8-56.2) |
| Q4 | 23.6% (23.2-24.0) | 13.4% (13.0-13.7) | 48.4% (47.7-49.1) | 27.4% (26.8-28.1) | 61.9% (61.0-62.9) | 39.4% (38.5-40.4) | 77.8% (76.8-78.8) | 52.0% (50.8-53.2) |
| Insurance Type | | | | | | | | |
| Commercial | 25.9% (25.6-26.1) | 15.8% (15.5-16.0) | 55.6% (55.2-56.1) | 33.9% (33.5-34.3) | 68.6% (68.1-69.2) | 46.2% (45.6-46.8) | 81.3% (80.7-81.8) | 56.7% (56.0-57.4) |
| Medicaid/ Replacement | 23.7% (22.8-24.7) | 12.6% (11.9-13.4) | 46.5% (44.9-48.1) | 24.7% (23.3-26.1) | 60.3% (57.9-62.6) | 37.4% (35.1-39.8) | 75.7% (73.1-78.2) | 48.9% (45.9-51.8) |
| Medicare/ Advantage | 21.8% (21.0-22.6) | 10.1% (9.5-10.7) | 37.1% (35.9-38.4) | 17.2% (16.2-18.1) | 48.0% (46.0-50.1) | 26.6% (24.8-28.5) | 75.0% (72.2-77.5) | 45.7% (42.6-48.7) |
| Self-pay | 16.5% (12.8-20.9) | 10.9% (7.9-14.7) | 43.4% (35.0-52.3) | 28.7% (21.4-37.3) | 52.5% (40.0-64.7) | 35.6% (24.6-48.3) | 67.5% (52.0-79.9) | 47.5% (32.9-62.5) |

Values are percentages (95% confidence interval) of patients achieving ≥5% or ≥10% total body weight loss (TBWL) at 6 and 12 months, among all ZEPBOUND initiators, ZEPBOUND initiators with a recorded weight, persistent users with a recorded weight, and persistent users at an initial FDA-labeled maintenance dose with a recorded weight, overall and stratified by sex, race/ethnicity, Social Vulnerability Index (SVI) quartile, and insurance type.

**Supplement Table 10. Adjusted Absolute Differences (in Percentage Points) in Prescribing Persistence, Maintenance Dose Attainment, and Weight Loss by Race and Ethnicity, Social Vulnerability Index, and Insurance Type.**

| | 12-Month Prescribing Persistence | FDA-Labeled Initial Maintenance Dose Attained by 12 Months | ≥5% Weight Loss at 12 Months |
|---|---|---|---|
| **Semaglutide** | | | |
| Race/Ethnicity | | | |
| NH White | Ref. (0) | Ref. (0) | Ref. (0) |
| Hispanic | -1.7 (-2.4, -1.1) | -0.9 (-1.4, -0.4) | -5.6 (-6.6, -4.5) |
| NH Black | 0.7 (0.2, 1.0) | -1.3 (-1.6, -0.9) | -8.8 (-9.4, -8.0) |
| NH Asian | 0.2 (-1.0, 1.4) | 1.1 (-0.3, 2.2) | -7.0 (-8.8, -4.9) |
| SVI | | | |
| Q1 | Ref. (0) | Ref. (0) | Ref. (0) |
| Q2 | -1.4 (-1.9, -0.9) | -1.0 (-1.4, -0.6) | -1.4 (-2.2, -0.5) |
| Q3 | -2.5 (-3.0, -2.0) | -2.4 (-2.8, -1.9) | -3.4 (-4.1, -2.7) |
| Q4 | -3.7 (-4.2, -3.2) | -4.3 (-4.8, -3.9) | -5.7 (-6.3, -4.9) |
| Insurance Type | | | |
| Commercial | Ref. (0) | Ref. (0) | Ref. (0) |
| Medicaid/Replacement | -2.7 (-3.3, -2.2) | -2.3 (-2.8, -1.9) | -2.3 (-3.1, -1.4) |
| Medicare/Advantage | -8.4 (-8.8, -7.9) | -14.5 (-14.9, -14.2) | -8.9 (-9.6, -8.2) |
| Self-pay | 2.8 (1.0, 5.7) | -11.0 (-12.7, -9.1) | -1.4 (-5.5, 2.2) |
| **Tirzepatide** | | | |
| Race/Ethnicity | | | |
| NH White | Ref. (0) | Ref. (0) | Ref. (0) |
| Hispanic | -3.0 (-3.8, -2.2) | -2.6 (-3.4, -1.9) | -4.9 (-6.0, -3.9) |
| NH Black | -3.2 (-3.8, -2.6) | -3.5 (-4.1, -2.9) | -9.2 (-10.2, -8.2) |
| NH Asian | 1.6 (0.3, 3.1) | 2.1 (0.5, 3.7) | -4.3 (-6.9, -2.0) |
| SVI | | | |
| Q1 | Ref. (0) | Ref. (0) | Ref. (0) |
| Q2 | -2.8 (-3.4, -2.1) | -3.1 (-3.7, -2.5) | -2.0 (-2.8, -1.1) |
| Q3 | -5.1 (-5.7, -4.5) | -5.7 (-6.4, -5.1) | -4.4 (-5.1, -3.5) |
| Q4 | -6.6 (-7.2, -6.0) | -7.7 (-8.4, -7.2) | -6.9 (-7.7, -6.0) |
| Insurance Type | | | |
| Commercial | Ref. (0) | Ref. (0) | Ref. (0) |
| Medicaid/Replacement | -9.3 (-10.2, -8.6) | -10.0 (-10.9, -9.3) | -7.1 (-8.7, -5.9) |
| Medicare/Advantage | -12.1 (-12.6, -11.5) | -17.5 (-18.2, -16.9) | -12.6 (-13.6, -11.7) |
| Self-pay | -1.0 (-4.5, 2.6) | 2.6 (-1.2, 6.3) | 0.9 (-4.5, 6.7) |

Values are adjusted absolute differences in percentage points (95% CI), estimated from multivariable logistic regression models fit separately for semaglutide and tirzepatide, with race/ethnicity, Social Vulnerability Index (SVI) quartile, and insurance type included together (mutually adjusted), and additionally adjusted for state and calendar month of index date. Adjusted absolute differences were estimated by marginal standardization relative to the indicated reference category (Ref., difference = 0 by definition); 95% confidence intervals were obtained by patient-level nonparametric bootstrap (200 replicates). NH denotes non-Hispanic.
